# Risk factors for long COVID: analyses of 10 longitudinal studies and electronic health records in the UK

**DOI:** 10.1101/2021.06.24.21259277

**Authors:** Ellen J. Thompson, Dylan M. Williams, Alex J. Walker, Ruth E. Mitchell, Claire L. Niedzwiedz, Tiffany C. Yang, Charlotte F. Huggins, Alex S. F. Kwong, Richard J. Silverwood, Giorgio Di Gessa, Ruth C.E. Bowyer, Kate Northstone, Bo Hou, Michael J. Green, Brian Dodgeon, Katie J. Doores, Emma L. Duncan, Frances M. K. Williams, OpenSAFELY Collaborative, Andrew Steptoe, David J. Porteous, Rosemary R. C. McEachan, Laurie Tomlinson, Ben Goldacre, Praveetha Patalay, George B. Ploubidis, Srinivasa Vittal Katikireddi, Kate Tilling, Christopher T. Rentsch, Nicholas J Timpson, Nishi Chaturvedi, Claire J. Steves

## Abstract

**Background:** The impact of long COVID is considerable, but risk factors are poorly characterised. We analysed symptom duration and risk factor from 10 longitudinal study (LS) samples and electronic healthcare records (EHR).

**Methods:** Samples: 6907 adults self-reporting COVID-19 infection from 48,901 participants in the UK LS, and 3,327 adults with COVID-19, were assigned a long COVID code from 1,199,812 individuals in primary care EHR. Outcomes for LS included symptom duration lasting 4+ weeks (long COVID) and 12+ weeks. Association with of age, sex, ethnicity, socioeconomic factors, smoking, general and mental health, overweight/obesity, diabetes, hypertension, hypercholesterolaemia, and asthma was assessed.

**Results:** In LS, symptoms impacted normal functioning for 12+ weeks in 1.2% (mean age 20 years) to 4.8% (mean age 63 y) of COVID-19 cases. Between 7.8% (mean age 28 y) and 17% (mean age 58 y) reported any symptoms for 12+ weeks, and greater proportions for 4+ weeks. Age was associated with a linear increased risk in long COVID between 20 and 70 years. Being female (LS: OR=1.49; 95%CI:1.24-1.79; EHR: OR=1.51 [1.41-1.61]), having poor pre-pandemic mental health (LS: OR=1.46 [1.17-1.83]; EHR: OR=1.57 [1.47-1.68]) and poor general health (LS: OR=1.62 [1.25-2.09]; EHR: OR=1.26; [1.18-1.35]) were associated with higher risk of long COVID. Individuals with asthma (LS: OR=1.32 [1.07-1.62]; EHR: OR=1.56 [1.46-1.67]), and overweight or obesity (LS: OR=1.25 [1.01-1.55]; EHR: OR=1.31 [1.21-1.42]) also had higher risk. Non-white ethnic minority groups had lower risk (LS: OR=0.32 [0.22-0.47]), a finding consistent in EHR. . Few participants had been hospitalised (0.8-5.2%).

**Conclusion:** Long COVID is associated with sociodemographic and pre-existing health factors. Further investigations into causality should inform strategies to address long COVID in the population.

## Introduction

SARS-CoV-2 infection can lead to sustained or recurrent multi-organ symptoms.^1–3^ Extended COVID-19 symptomatology over weeks to months has been termed ‘long COVID’.^4^ The UK’s National Institute for Health Care and Excellence (NICE) distinguishes acute COVID-19 (AC; lasting <4 weeks), ongoing symptomatic COVID-19 (OSC; 4-12 weeks), and post-COVID-19 syndrome (PCS; >12 weeks), with the latter two both considered ‘long COVID’.^1^ Long COVID prevalence estimates range from 13.3% in highly selected, community-based survey respondents with test-confirmed COVID-19, to at least 71% among those hospitalised by the infection.^5–7^ Given the scale of the pandemic, even a low proportion of individuals with long COVID will generate a major burden of enduring illness.^8^

Targeting appropriate support and research first entails identification of putative risk factors for the disease. Current understanding of long COVID risk factors and their frequency remains poor, impeding mechanistic understanding and, intervention and evidence-based service planning. Accurate risk estimates require large generalisable samples with comprehensive measures of pre-pandemic characteristics. UK national primary care records (EHR), covering >95% of the population, afford one data source, but are limited to those seeking care, obtaining diagnosis long COVID, and gaining a subsequent diagnostic code. Established population-based longitudinal studies (LS), overcome these limitations by collecting data from participants regardless of healthcare attendance, and benefit from measures of pre-pandemic characteristics. While individual LS are relatively small, combining data from multiple studies yield large sample sizes. Triangulation of findings with equivalent results from EHR can further compensate for different limitations and biases.

To meet clinical and policy needs, we identified individuals with OSC and PCS (long COVID) in: 1) a consortium of population-based LS which captured coordinated repeat questionnaire data on COVID-19 using harmonised measures from the <u>Wellcome Trust’s Covid-19 Questionnaire</u>, and 2) the OpenSAFELY dataset of primary care records (https://www.opensafely.org/). Here, we report the frequency of long COVID among individuals with suspected and test-confirmed COVID-19 and examined associations with sociodemographic and pre-pandemic health risk factors.

## Methods

### Design

The UK National Core Studies – Longitudinal Health and Wellbeing programme (https://www.ucl.ac.uk/covid-19-longitudinal-health-wellbeing/) combines data from multiple UK population-based LS and electronic health records (EHR) to answer pandemic-relevant questions. In this analysis we pooled results from parallel analyses within individual LS, then compared with population-based findings from EHR capturing individuals who actively sought healthcare.

### Sample

#### Longitudinal Studies (LS)

Data were drawn from 10 UK LS that had conducted surveys before and during the COVID-19 pandemic comprising five age-homogenous cohorts: the Millennium Cohort Study (MCS); the Avon Longitudinal Study of Parents and Children (ALSPAC (generation 1, “G1”)); Next Steps (NS); the 1970 British Cohort Study (BCS); and the National Child Development Study (NCDS), and five age-heterogeneous samples were included: the Born in Bradford study (BIB); Understanding Society (USOC); Generation Scotland: the Scottish Family Health Study (GS); the parents of the ALSPAC-G1 cohort, whom we refer to as ALSPAC-G0; and the UK Adult Twin Registry (TwinsUK). Study details and references are shown in Supplementary Table 1. Minimum inclusion criteria were pre-pandemic health measures, age, sex, ethnicity plus self-reported COVID-19, and self-reported duration of COVID-19 symptoms. Ethics statements are presented in Supplementary Table 2.

#### Electronic Health Records (EHR)

Working on behalf of NHS England, we conducted a population-based cohort study to measure long COVID recording in electronic health record (EHR) data from primary care practices using TPP SystmOne software, linked to Secondary Uses Service (SUS) data (containing hospital records) through OpenSAFELY (https://www.opensafely.org/). This is a data analysis platform developed on behalf of NHS England during the COVID-19 pandemic to allow near real-time analysis of pseudonymised primary care records within the EHR vendor’s highly secure data environment to protect patient privacy. Details on Information Governance for the OpenSAFELY platform can be found in the Supplementary Information 1. From a population of all people alive and registered with a general practice on 1 December 2020, we selected all patients who had evidence of a COVID-19 related code, either: positive SARS-CoV-2 testing, being hospitalised with an associated COVID diagnostic code, or having a recorded diagnostic code for COVID in primary care.

#### Measures

##### Outcomes: COVID-19 and long COVID definitions

LS: COVID-19 cases were defined by self-report, including testing confirmation and health care professional diagnosis (see Supplementary File 1 for full details of the questions and coding used within each study). Long COVID was defined as per NICE as either OSC or PCS using self-reported symptom duration.^1^ Based on these categories, we defined two primary outcomes: i) symptoms lasting 4+ weeks (combining OSC and PCS, symptoms lasting 0-4 weeks as reference), and ii) symptoms lasting 12+ weeks (PCS specifically, symptoms lasting 0-12 weeks as reference). Some studies recorded duration of symptoms of any severity, whereas others referred only to symptoms which impacted daily function. In addition, two studies derived alternate estimates of long COVID based on individual symptom counts lasting more than 4 or 12 weeks over at least six months (BiB, TwinsUK) (Supplementary Information 2).

EHR: Any record of long COVID in the primary care record was coded as a binary variable. This was defined using a list of 15 UK SNOMED codes, categorised as diagnostic (2 codes), referral (3) and assessment (10) codes. SNOMED is an international structured clinical coding system for use in EHR. The outcome was measured between the study start date (2020-02-01) and the end date (2021-05-09).

#### Exposures

##### Sociodemographic factors

All studies included age, sex, ethnicity (white or non-white ethnic minority, where available) and Index of Multiple Deprivation (IMD; divided into quintiles). LS included additional measures of socioeconomic position: education (degree, no degree), and occupational class of own current/recent employment (Supplementary file 1). EHR also included geographic region.^9^

#### Mental health

LS: Pre-pandemic measures using validated continuous scales of anxiety and depression symptoms dichotomised using established cut-offs to indicate distress (see Supplementary file 1).

EHR: Evidence of a pre-existing mental health condition was defined using prior codes for one of: psychosis; schizophrenia; bipolar disorder; or depression.

#### Self-rated general health

LS: Pre-pandemic self-rating on a 5-point scale dichotomised to compare excellent-good health (categories 1-3) with fair-poor health (categories 4-5).

#### Overweight and obesity

LS: Body mass index (BMI; kg/m^2^) obtained prior to the pandemic, coded to compare a BMI between 0-24.9 (having underweight/normal weight) against a BMI of >=25 (overweight/obesity).

EHR: Categorised as having or not having obesity using the most recent BMI measurement, with those having obesity further classified into having Obese I (BMI 30-34.9), Obese II (BMI 35-39.9), or Obese III (BMI 40+).

#### Health conditions

LS: Pre-pandemic self-report of asthma, diabetes, hypertension, and high cholesterol status. EHR: A previous code six months to five years before March 2020 for one or more of: diabetes; cancer; haematological cancer; asthma; chronic respiratory disease; chronic cardiac disease; chronic liver disease; stroke or dementia; other neurological condition; organ transplant; dysplasia; rheumatoid arthritis, systemic lupus erythematosus or psoriasis; or other immunosuppressive conditions. Those with no relevant code for a condition were assumed not to have that condition. Number of conditions were categorised into “0”, “1”, and “2 or more”.

#### Statistical analysis: LS

Main analyses were conducted in studies with a direct self-reported measure of COVID-19 symptom length. Associations between each factor and both long COVID outcomes (long COVID and PCS) were assessed in separate logistic regression models within each study. We adjusted for a minimal set of confounders across all studies, where relevant: age (adjusted as a continuous variable), sex, and ethnicity. We report odds ratios (ORs) and 95% confidence intervals (CIs).

We modelled the relationship of age with long COVID risk in two ways. First, in age-heterogeneous samples we compared long COVID risk within pre-defined age categories (see Supplementary figures 1 and 2). Second, in a subset of LS birth cohorts with participants of near-identical ages and who were issued fully harmonised long COVID questionnaires (MCS, NS, BCS70 and NCDS), we analysed the trend in absolute risk of long COVID with increasing age between studies using meta-regression.

Attrition and survey design were addressed by weighting estimates to be representative of their target population in each LS (weights were not available for BiB and TwinsUK).

To synthesise effect sizes across studies, fixed-effect meta-analysis with restricted maximum likelihood was carried out and repeated with random-effects modelling for comparison. The *I*^*2*^ statistic was used to report heterogeneity between estimates.

##### Sensitivity analyses

To mitigate index event bias,^10^ inverse probability weights (IPW) were derived for risk of COVID-19. These were derived in each LS separately but following a common approach used previously.^11^ Derived weights were then applied in all analysis models as a sensitivity check (see Supplementary Information 3).

For studies in which we were able to verify SARS-CoV-2 infection (TwinsUK and ALSPAC-G0 and -G1), analyses were replicated on a sub-sample of those who had positive polymerase chain reaction (PCR) obtained through linkage to testing data and/or lateral flow antibody testing (ALSPAC) and enzyme-linked immunosorbent assay (ELISA) (TwinsUK)^12^ confirming viral exposure.

#### Statistical Analysis: EHR

We conducted logistic regression to assess whether GP-recorded long COVID was associated with each sociodemographic or pre-pandemic health characteristic. We adjusted for the same set of confounders as used in the LS analyses: age (as categorical variable), sex, ethnicity.

In further analyses of age as a risk factor for long COVID in the EHR data, we assigned individuals within 10-year categories an age at the midpoint of each group, then assessed the trend in long COVID frequency with age using linear and non-linear meta-regression.

All code for the OpenSAFELY platform for data management, analysis and secure code execution is shared for review and re-use under open licenses at https://github.com/opensafely. All codelists (describing the definition of the conditions) and the code for data management and analysis is shared for scientific review and re-use under open licenses on GitHub https://github.com/opensafely/long-covid-historical-health

## Results

### Long COVID descriptives

Of 48,901 individuals surveyed in LS, 6907 (14.12%) self-reported suspected or confirmed COVID-19. (Table 1 and Supplementary Table 3).

**Table 1:**
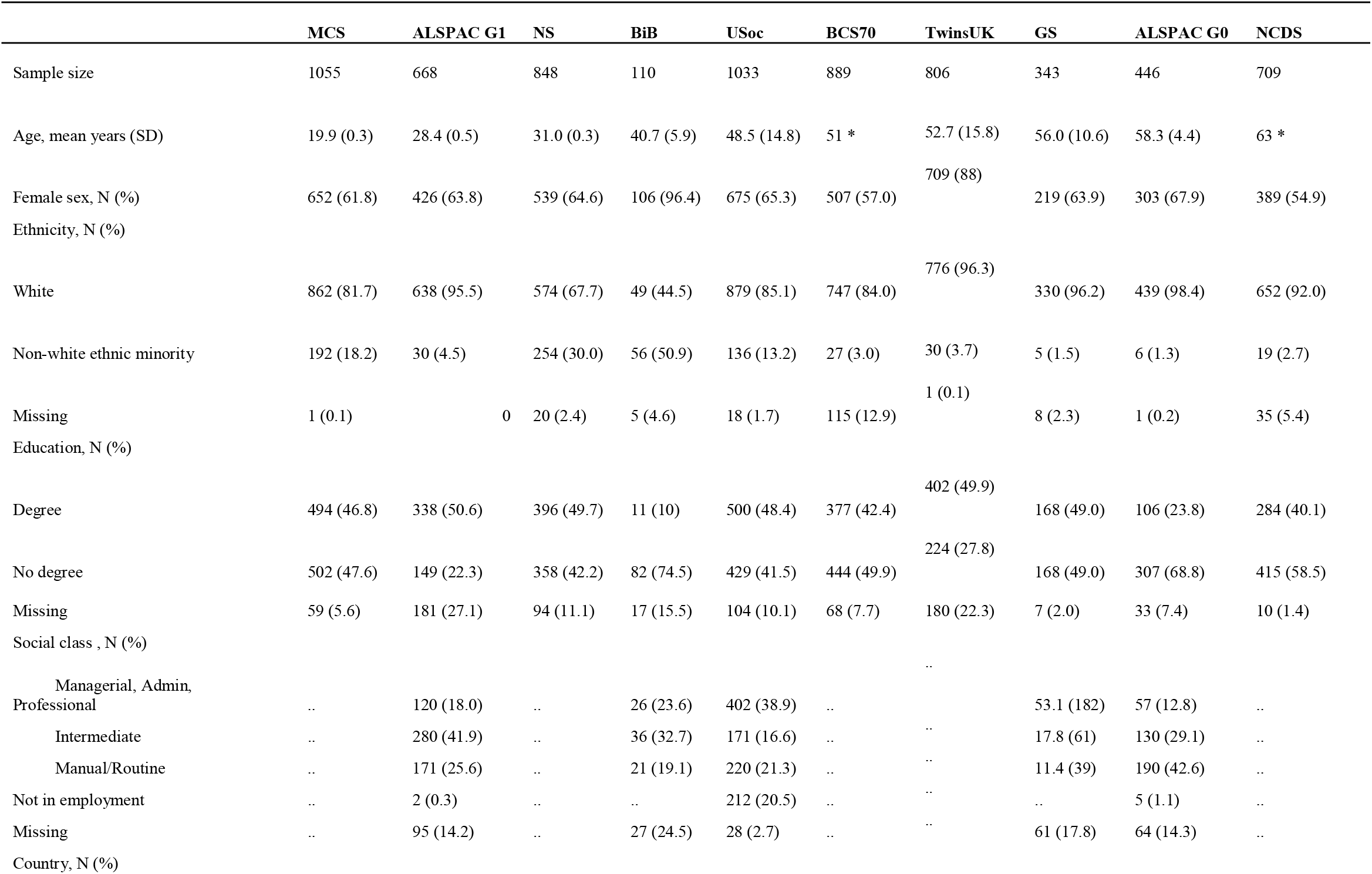
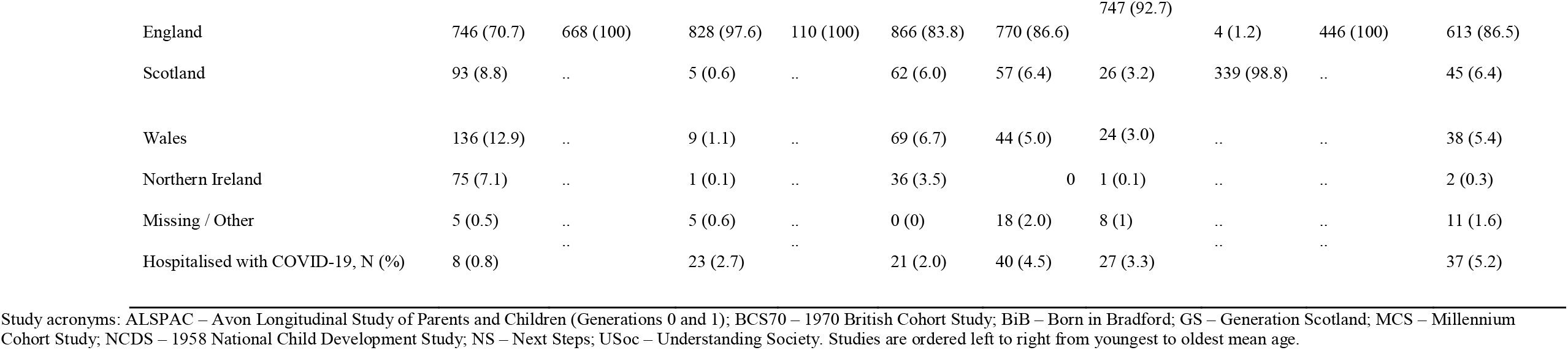
Characteristics of the analytic samples from the longitudinal studies (self-reported COVID-19 cases with data on duration of symptoms)

In LS, reporting of OSC (symptoms 4-12 weeks), ranged between 14.5-18.1%, and for PCS (symptoms 12+ weeks) 7.8-17%. When restricted to symptoms limiting day-to-day function, proportions were lower, ranging from 3.0-13.7% for OSC, and 1.2-4.8% for PCS (see Table 2). Figures varied considerably within LS comparing self-reported confirmed and suspected cases (see Supplementary Table 4). However, when stratifying results to those who had a positive SARS-CoV-2 antibodies/PCR, reporting of OSC ranged from 8.8 to 20%, and PCS ranged from 11 to 20% (see Supplementary Table 5).

**Table 2:** Symptoms duration among self-reported COVID-19 cases in the longitudinal studies

| Study | COVID-19 cases with symptom duration data | Mean age | Duration of symptoms, N (%) |  |  |
| --- | --- | --- | --- | --- | --- |
|  |  |  | Acute (0-4 weeks) | Ongoing symptomatic COVID-19 (4-12 weeks) | Post COVID-19 syndrome (12+ weeks) |
| *Studies ascertaining long COVID of any severity* |  |  |  |  |  |
| ALSPAC G1 | 668 | 28.4 | 519 (77.7) | 97 (14.5) | 52 (7.8) |
| USoc | 1033 | 48.5 | 742 (71.8) | 182 (17.6) | 109 (10.6) |
| TwinsUK | 806 | 52.7 | 579 (71.8) | 146 (18.1) | 81 (10) |
| GS | 335 | 56.0 | 224 (66.9) | 54 (16.1) | 57 (17.0) |
| ALSPAC G0 | 446 | 58.3 | 302 (67.7) | 68 (15.2) | 76 (17.0) |
| *Studies ascertaining severe long COVID only \** |  |  |  |  |  |
| MCS | 1055 | 19.9 | 1010 (95.7) | 32 (3.0) | 13 (1.2) |
| Next Steps | 848 | 31.0 | 773 (91.2) | 51 (6.0) | 24 (2.8) |
| BCS70 | 889 | 51.0 | 757 (85.2) | 84 (9.5) | 48 (5.4) |
| NCDS | 709 | 63.0 | 578 (81.5) | 97 (13.7) | 34 (4.8) |
| *Studies ascertaining long COVID by monthly symptom reporting \*\** |  |  |  |  |  |
| BiB | 110 | 40.7 | 40 (36.4) | 26 (22.7) | 46 (40.9) |
| TwinsUK | 953 | 54 | 272 (28.5) | 246 (25.8) | 435 (45.6) |

Study acronyms: ALSPAC – Avon Longitudinal Study of Parents and Children (Generations 0 and 1); BCS70 – 1970 British Cohort Study; BiB – Born in Bradford; GS – Generation Scotland; MCS – Millennium Cohort Study; NCDS – 1958 National Child Development Study; NS – Next Steps; USoc – Understanding Society.
**Footnotes:** Studies are ordered from youngest to oldest mean age within categories of method of long COVID ascertainment
\* Questionnaires in these four cohorts asked respondents to report duration for which COVID-19 symptoms impeded normal function, rather than simply the duration of any symptoms (however mild) as in other studies. Hence proportions reporting long COVID in them are expected to be lower when compared to other cohorts with similar characteristics
\*\* Based on symptom-counting approach over months, rather than self-reported duration of symptoms as in all other cohorts, which yields higher proportions of individuals being designated long COVID categories

Two studies used an individual symptoms approach (recorded retrospectively over several months) to ascertain symptom duration among individuals with COVID-19. Using this approach, reporting of 4-12 weeks ranged between 22.7-25.8% and 12+ weeks ranged between 40.9-45.6% (Table 2). However, high proportions were also found in those who had not had COVID-19 with this ascertainment method (12+ weeks, 28.8%, 4-12 weeks, 21.8%, and 0-4 weeks, 17.8%, Supplementary Table 6). Therefore, data for long COVID ascertained in this way were not taken forward to risk factor analysis due to uncertainty that they met NICE criteria, which require that symptoms are “not explained by an alternative diagnosis”.

In the EHR, within 1,199,812 individuals with any acute COVID-19 code, 3327 individuals also had a recorded long COVID code, constituting 0.27% of COVID-19 cases.

#### Age and long COVID

Symptom reporting increased with age to age ∼70 years in the LS, for both OSC and PCS. Using age homogenous LS, we observe a 3.02% (95% CI: 1.86-4.17) rise per decade with age for functionally limiting symptoms lasting 4+ weeks, and 0.68% (95% CI: -0.15-1.51) for functionally limiting PCS between 20-63 years of age (Figure 1). In the age-heterogeneous LS, increasing trends in risk of symptoms lasting both 4+ weeks and 12+ weeks with higher age were observed across participants ranging from young adulthood to approximately 70 years (Supplementary Figures 1 and 2) OpenSAFELY, showed an inverted U-shaped association of long COVID with age, (Supplementary Figure 1), with highest rates in those aged 45-54, and 55-69 years. People aged 80 and above had no higher risks compared to the reference group aged 18-24 years.

**Figure 1:**
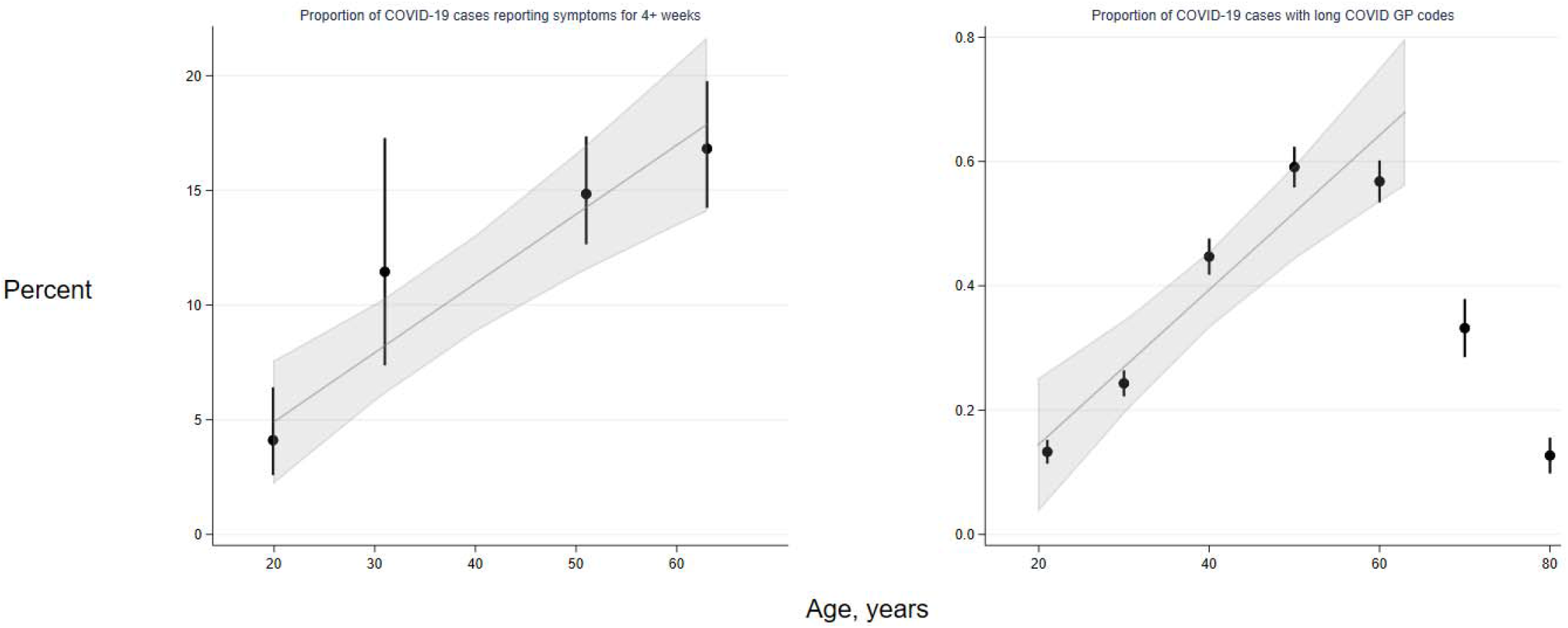
Trends in long COVID frequency among COVID-19 cases by age, in four age-homogeneous LS (left) and EHRs (right) Legend: Left -- in four longitudinal studies where participants are of near-identical ages (the cohorts MCS, NS, BCS70 and NCDS), proportions reporting symptom length of four or more weeks in COVID-19 cases were ascertained from questionnaire responses. Right -- in OpenSAFELY, proportions represent individuals within 10-year age categories (with estimates grouped at the mid-point of each category) who have long COVID codes in GP records, hence the proportions are substantially lower than in the corresponding cohort data. Trend lines and 95% confidence interval shading represent absolute differences in long COVID frequencies with increasing age, estimated by linear meta-regression of data from the four cohorts and from 18 to 70 year olds in OpenSAFELY (data from older individuals were not modelled; refer to results text for further explanation).

There was a linear increase of absolute risk of long COVID of 0.12% per decade (95% CI: 0.08-0.17) between 18 and 70 years, aligning with LS results (Figure 1, right panel).

#### Socio-demographic factors and long COVID

Figure 2 shows pooled associations in LS (10 cohorts, n=6907 cases) between other sociodemographic and health factors and each binary long COVID outcome (see Supplementary Figures 3 to 6 for study-level results). Females had higher risk of both long COVID outcomes (4+ weeks: OR=1.49; 95%CI: 1.24-1.79; 12+ weeks: OR=1.60; 95%CI: 1.23-2.07). No clear evidence was found for individuals of non-white ethnicity (compared to individuals of white ethnicity) having differential risk of OSC and PCS combined (OR for symptoms lasting 4+ weeks =0.80; 95%CI: 0.54-1.19). Non-white ethnicity was associated with lower risk of PCS specifically (OR=0.32; 95%CI: 0.22-0.47) after meta-analysis, but study-level findings displayed a high degree of heterogeneity (*I*^*2*^=75%, *P*<0.001; Supplementary figure 5). Across LS, no strong evidence was found for association of IMD with either outcome. Having not attained a degree from higher education was associated with lower risk of PCS (OR: 0.73; 95% CI: 0.57-0.94), but not with OSC and PCS in combination (OR: 0.95: 95% CI: 0.80-1.14).

**Figure 2:**
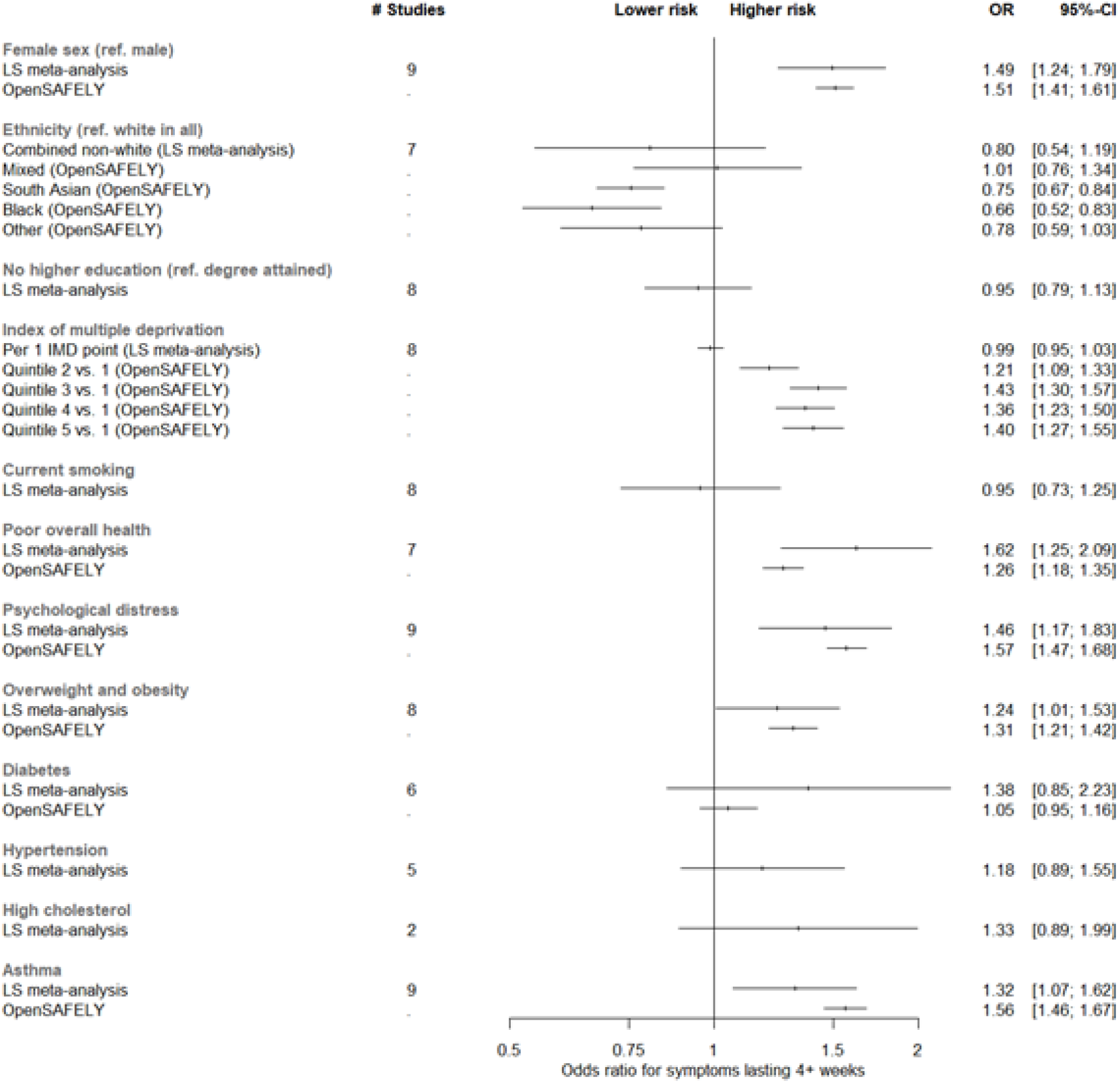
Risk factors associated with long COVID from meta-analyses of LS findings alongside corresponding analyses from EHRs Legend: All associations were adjusted for age and sex, except where redundant. In all instances where it was possible to derive results from both meta-analyses of longitudinal studies and analysis of EHRs, the corresponding results are plotted side-by-side for comparison. The outcome used for longitudinal study fixed-effect meta-analysis estimates presented here was symptoms lasting for 4+ weeks, and the outcome in EHRs was any reporting of a long COVID read code in GP records (regardless of duration of symptoms). Full study-level results, heterogeneity statistics and random-effect estimates for the longitudinal study meta-analyses are presented in supplemental figures 3 and 4. The equivalent meta-analyses of longitudinal study data where symptom duration of 12+ weeks was instead used as the outcome are depicted in supplemental figures 5 and 6. ‘Poor overall health’ represents the self-rated health exposure in the LS meta-analysis, and comorbidities in OpenSAFELY. The outcome ‘Overweight and obesity’ represents combined BMI categories over 25 in the LS, and solely individuals with BMI 30-34.9 in OpenSAFELY.

In EHR, females had higher risk of long COVID than males (OR=1.51; 95%CI:1.41-1.61), while odds were lower in individuals of South Asian (compared to (OR=0.75; 95%CI:0.67-0.84) or black ethnicity, relative to white ethnicity (OR=0.66; 95%CI:0.52-0.83) (Table 3 and Figure 2). Individuals living in areas with the least deprivation had higher odds of a long COVID code compared to those in the most deprived IMD quintile (Figure 2).

**Table 3:**
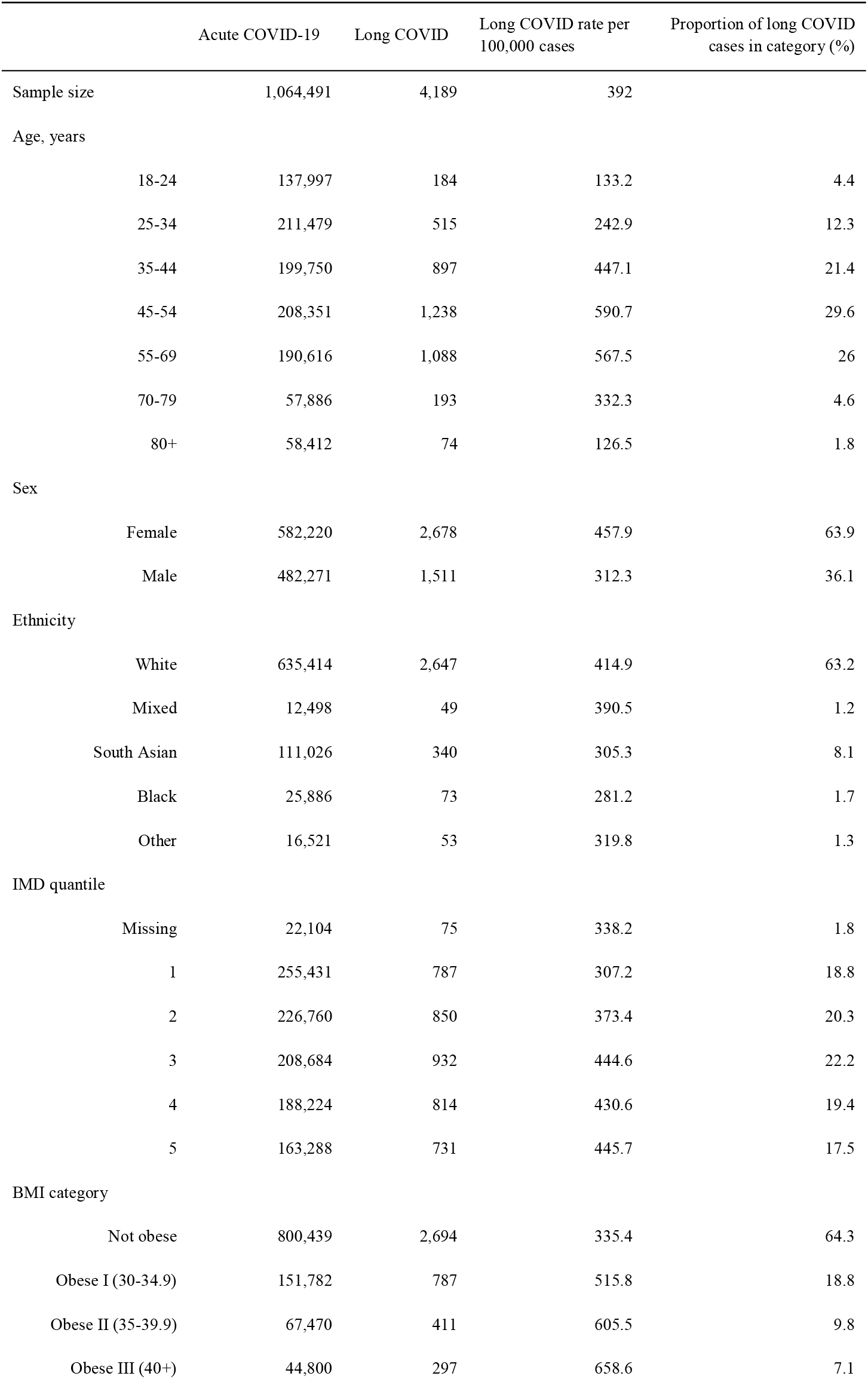
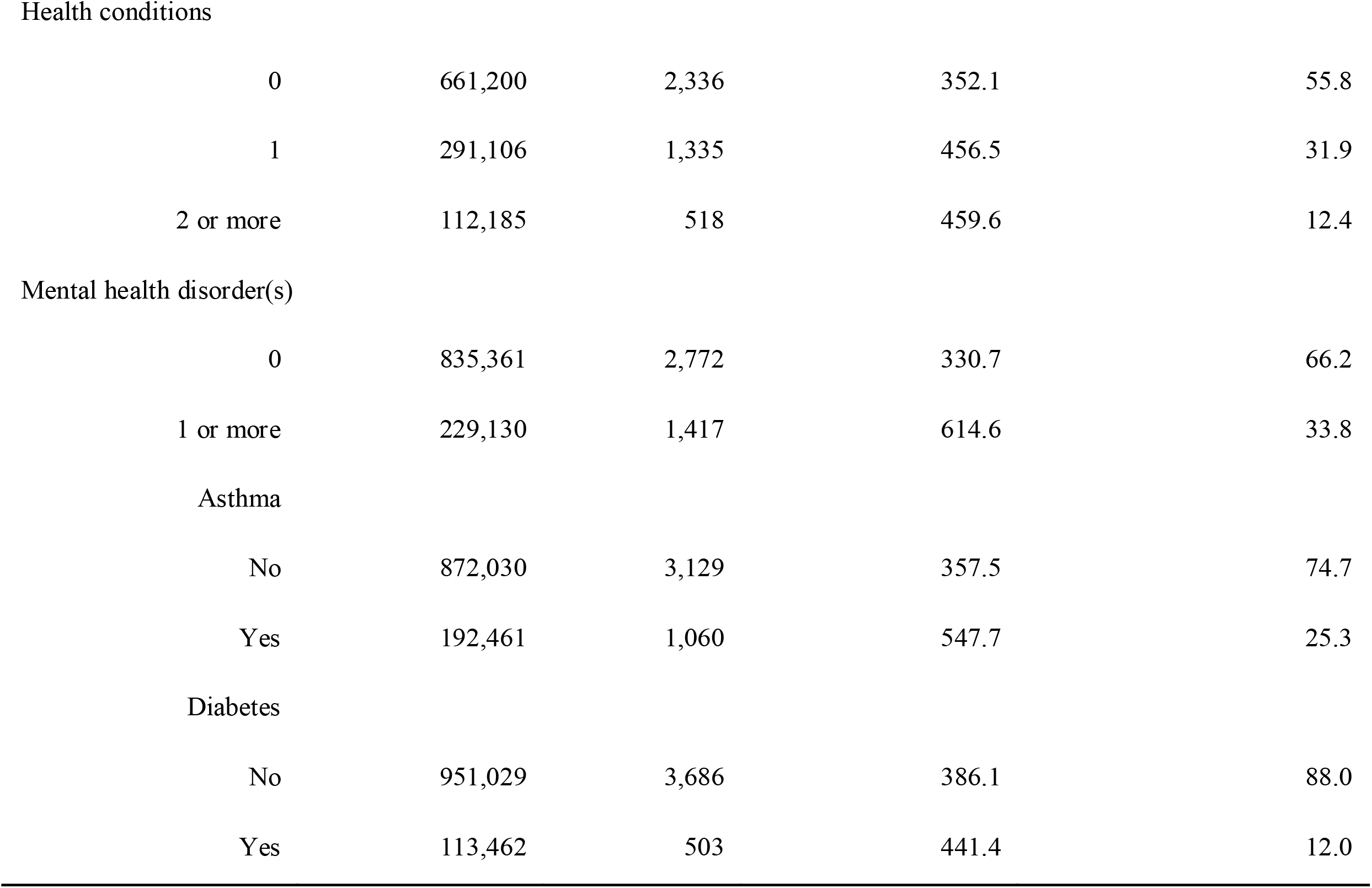
Characteristics of individuals reported to have had COVID-19 and long COVID by general practitioners in OpenSAFELY

|  | Acute COVID-19 | Long COVID | Long COVID rate per 100,000 cases | Proportion of long COVID cases in category (%) |
| --- | --- | --- | --- | --- |
| Sample size | 1,064,491 | 4,189 | 392 |  |
| Age, years |  |  |  |  |
| 18-24 | 137,997 | 184 | 133.2 | 4.4 |
| 25-34 | 211,479 | 515 | 242.9 | 12.3 |
| 35-44 | 199,750 | 897 | 447.1 | 21.4 |
| 45-54 | 208,351 | 1,238 | 590.7 | 29.6 |
| 55-69 | 190,616 | 1,088 | 567.5 | 26 |
| 70-79 | 57,886 | 193 | 332.3 | 4.6 |
| 80+ | 58,412 | 74 | 126.5 | 1.8 |
| Sex |  |  |  |  |
| Female | 582,220 | 2,678 | 457.9 | 63.9 |
| Male | 482,271 | 1,511 | 312.3 | 36.1 |
| Ethnicity |  |  |  |  |
| White | 635,414 | 2,647 | 414.9 | 63.2 |
| Mixed | 12,498 | 49 | 390.5 | 1.2 |
| South Asian | 111,026 | 340 | 305.3 | 8.1 |
| Black | 25,886 | 73 | 281.2 | 1.7 |
| Other | 16,521 | 53 | 319.8 | 1.3 |
| IMD quantile |  |  |  |  |
| Missing | 22,104 | 75 | 338.2 | 1.8 |
| 1 | 255,431 | 787 | 307.2 | 18.8 |
| 2 | 226,760 | 850 | 373.4 | 20.3 |
| 3 | 208,684 | 932 | 444.6 | 22.2 |
| 4 | 188,224 | 814 | 430.6 | 19.4 |
| 5 | 163,288 | 731 | 445.7 | 17.5 |
| BMI category |  |  |  |  |
| Not obese | 800,439 | 2,694 | 335.4 | 64.3 |
| Obese I (30-34.9) | 151,782 | 787 | 515.8 | 18.8 |
| Obese II (35-39.9) | 67,470 | 411 | 605.5 | 9.8 |
| Obese III (40+) | 44,800 | 297 | 658.6 | 7.1 |
| Health conditions |  |  |  |  |
| 0 | 661,200 | 2,336 | 352.1 | 55.8 |
| 1 | 291,106 | 1,335 | 456.5 | 31.9 |
| 2 or more | 112,185 | 518 | 459.6 | 12.4 |
| Mental health disorder(s) |  |  |  |  |
| 0 | 835,361 | 2,772 | 330.7 | 66.2 |
| 1 or more | 229,130 | 1,417 | 614.6 | 33.8 |
| Asthma |  |  |  |  |
| No | 872,030 | 3,129 | 357.5 | 74.7 |
| Yes | 192,461 | 1,060 | 547.7 | 25.3 |
| Diabetes |  |  |  |  |
| No | 951,029 | 3,686 | 386.1 | 88.0 |
| Yes | 113,462 | 503 | 441.4 | 12.0 |

#### Health factors and long COVID

In LS, those with poor or fair pre-pandemic self-reported general health had greater risk of both long COVID outcomes (4+ weeks: OR=1.62; 95%CI: 1.25-2.09; 12+ weeks: OR=1.66; 95%CI: 1.14-2.40). Greater pre-pandemic psychological distress was also associated with higher risk of both long COVID outcomes (4+ weeks: OR=1.45; 95%CI: 1.16-1.82; 12+ weeks: OR=1.58; 95%CI: 1.15-2.17). No strong evidence was observed for a linear association of BMI with either outcome, while overweight/obesity was associated with increased odds of symptoms lasting for 4+ weeks (OR= 1.24; 95%CI: 1.01-1.53) but not with PCS specifically (OR 0.95, 95% CI: 0.70-1.28). Associations were not found for diabetes, hypertension, or high cholesterol with either outcome, although modest point estimates were on the side of higher long COVID risk in several instances (**Supplementary figures 4 and 6)**. Asthma was the only specific medical condition associated with increased odds of having symptoms for 4+ weeks (OR=1.31; 95%CI: 1.06-1.62), although the association with PCS specifically was closer to the null (OR=1.12; 95%CI: 0.80-1.58).

In EHR, increased odds of having a long COVID code was seen in individuals with pre-existing comorbidities (OR=1.26; 95%CI:1.18-1.35) and psychiatric conditions (OR=1.57; 95%CI:1.47-1.68). An increased risk was observed in individuals with a pre-pandemic diagnosis of asthma (OR=1.56; 95%CI:1.46-1.67) and overweight and obesity (OR=1.31, 95%CI:1.21-1.42). No increase in risk was observed for diabetes.

##### Sensitivity analyses

In LS, when including IPWs for risk of COVID-19 status, all identified associations persisted and, in some instances, associations increased slightly in magnitude **(Supplementary figures 7 to 10)**. Notably hypercholesterolaemia was associated with both long COVID outcomes in the LS meta-analyses weighted for probability of reporting COVID-19.

## Discussion

In parallel analyses of 10 population-based longitudinal studies and 1.2 million primary care EHRs, we observed varying proportions of adults with COVID-19 who had long COVID depending on the age of study members and the degree of functional impact. While just 0.3% of COVID-19 cases had long COVID codes in primary care, up to 17% of adult COVID-19 cases in midlife reported symptoms attributed to COVID-19 for more than 12 weeks in longitudinal studies. Clear associations between long COVID risk and sociodemographic characteristics (older age, female sex, white ethnicity) and antecedent health factors (poor mental and general health, asthma) were also established.

Recent reports of the frequency of long COVID vary, with the REACT-2 study reporting 14.8% of COVID-19 cases with 3 or more symptoms persisting for 12+ weeks, and 11.5% of COVID-19 cases with 3 or more enduring symptoms affected their daily lives.^13^ These estimates are significantly higher than our estimates of functionally limiting PCS (1.2 to 4.8%, according to age). However, as detailed above, this definition diverges from the NICE definition which requires symptoms not to be attributable to an alternative cause.

Using a similar symptom counting approach in our study, we found that the proportion of OSC and PCS were largely consistent with other population-based studies.^4,13,14^ However, high rates of symptom reporting were also found in those without COVID-19, thus estimates using this approach should be treated with caution. Notable discordance in these proportions would yield very different prevalence estimates for the number of people in the UK population presumably requiring care for debilitating long COVID, with REACT-2 estimating 1.7% of the English population, and the ONS estimating approximately 0.3% of the UK population.^13,15^ Several reasons could explain disparities in observed proportions with the most severe long COVID, including estimates for England vs. the UK as a whole, questionnaire wording, basing estimates on test-validated versus self-reported COVID-19 cases, and representativeness (REACT-2’s response rates being 26 to 29%; ONS reporting 51% for its May 2021 survey; and in the most recent LS surveys, response rates for studies reporting of functionally limiting PCS ranged from 33 to 58.5%).

The lower reporting of long COVID in primary care compared to our LS data and other studies suggest that only a minority of people with long COVID seek care and/or subsequently receive a code. Diagnostic codes for long COVID have only recently been instituted and uptake by primary care practitioners has not been uniform.^16^ The analyses here are based on practices that use TPP SystmOne software and is therefore limited to England, and we note that these practices had a 2- to 3-fold lower rate of long COVID recording than those that use EMIS software.^16^

Despite definition differences in primary care versus LS, several risk factor associations were consistent between various LS and in EHR. In both LS and EHR, long COVID reporting by any definition increased with age. Unlike risk of severe COVID-19, this appeared to be linear (not exponential) across most adult age groups. In individuals aged over 70 we observed a sharp decline in long COVID risk in most LS and the EHR data. This decline in older age has been observed in other studies,^4,17,18^ and may be spurious due to selective competing risk of mortality, non-response bias, lower symptom reporting in older adults, misattribution of long COVID to other illness, or a combination of these factors. The findings that long COVID was 50% higher in women than men is consistent with reports from most^4,17,19–22^ but not all previous studies.^4,18^ We found some evidence of higher long COVID reporting among individuals of white ethnicity and of higher educational attainment, which was unexpected given the common associations of these characteristics with lower morbidity more generally. While we found no strong evidence for a relationship between area-level socioeconomic status in LS, in primary care EHR there was also an apparent gradient of higher risk in individuals from the least deprived areas. These associations could reflect unmet need in medical care for those who live in socioeconomically deprived areas or circumstances. However, these results contrast with two studies reporting null findings for ethnicity and socioeconomic status in relation to long COVID from other countries,^18,23^ and the ONS and REACT-2 surveys reported similar associations for ethnicity, but opposite associations in long COVID reporting by deprivation scales, to that which we observed in EHR.^13^

A greater risk of long COVID related to adverse prior mental health, has been reported elsewhere,^23^ but pre-pandemic general health has not previously been highlighted as a risk factor.^17,20,24^ Excess risk of long COVID in association with asthma across cohorts and primary care records resolves conflicting and limited findings,^4,17,23^ and supports a focus on asthma as a high-risk condition, for example by investigating whether immune processes involved in asthma or respiratory complications influence long COVID development. Findings for overweight/obesity were suggestive of an increased risk, again helping to resolve some previous uncertainty.^4,17,20,25^ No other cardiometabolic risk factors were clearly associated, consistent with past studies.^4,20,23,25^

A major strength of this research was the coordinated investigation of long COVID in multiple LS and EHR, each with differing bias, study designs, target populations, and selection and attrition processes. Consistent findings emerging from these sources add reliability. We used population-based resources to increase the representativeness of findings to long COVID in the community. Unlike newly established studies which have collected exposure data during the pandemic, the long-running data collections in both LS and EHR allowed us to study *prospective* associations of risk factors with long COVID, meaning results will not have arisen from reverse causation, nor will exposure definitions have been influenced by recall bias. Rich antecedent data also allowed us to run a range of sensitivity analyses to re-weight our results for non-response (reducing the bias from selection into samples). We also flag important limitations, principally that our data are observational, and we cannot draw causal conclusions on the role of risk factors in long COVID development, and that whilst we attempted to address both selection into our samples from study attrition and selecting upon COVID-19 case status (which can induce index event bias,^10^ there remains the possibility that potential bias has influenced association estimates. Finally, not all studies had test confirmation of COVID-19 status, and some individuals may have misattributed persistent symptoms to other conditions.

### Implications

The stark variability in proportions of COVID-19 cases with persistent symptoms is clear from our comparison of methods of ascertaining long COVID. Representative population-based studies will need to provide ongoing estimates across the spectrum of functional limitation to help plan appropriate provision of healthcare. Our data suggest that revisions of diagnostic criteria within primary care may be appropriate, particularly for demographic groups which are less in touch with healthcare services. Although causal inferences cannot be drawn from these data, our findings justify further investigations into the role of sex difference, age related change, and/or immunity and respiratory health in development of long COVID. Older working individuals, with high levels of comorbidity, may particularly require support.

## Data Availability

Data for NCDS (SN 6137), BCS70 (SN 8547), Next Steps (SN 5545), MCS (SN 8682) and all four COVID-19 surveys (SN 8658) are available through the UK Data Service. NSHD data are available on request to the NSHD Data Sharing Committee. Interested researchers can apply to access the NSHD data via a standard application procedure. Data requests should be submitted to; further details can be found at http://www.nshd.mrc.ac.uk/data.aspx. doi:10.5522/NSHD/Q101; doi:10.5522/NSHD/Q10
ALSPAC data is available to researchers through an online proposal system. Information regarding access can be found on the ALSPAC website (http://www.bristol.ac.uk/media-library/sites/alspac/documents/researchers/data-access/ALSPAC_Access_Policy.pdf).
Data from the various BiB family studies are available to researchers; see the study website for information on how to access data (https://borninbradford.nhs.uk/research/how-to-access-data/).
All data for Understanding Society  are available through the UK Data Service (SN 6614 and SN 8644).
Access to data is approved by the Generation Scotland Access Committee. See https://www.ed.ac.uk/generation-scotland/for-researchers/access or for further details.
The TwinsUK Resource Executive Committee (TREC) oversees management, data sharing and collaborations involving the TwinsUK registry (for further details see https://twinsuk.ac.uk/resources-for-researchers/access-our-data/).

## People acknowledgments

OpenSAFELY Collaborative Group: Alex J Walker, Brian MacKenna, Peter Inglesby, Christopher T Rentsch, Helen J Curtis, Caroline E Morton, Jessica Morley, Amir Mehrkar, Seb Bacon, George Hickman, Chris Bates, Richard Croker, David Evans, Tom Ward, Jonathan Cockburn, Simon Davy, Krishnan Bhaskaran, Anna Schultze, Elizabeth J Williamson, William J Hulme, Helen I McDonald, Laurie Tomlinson, Rohini Mathur, Rosalind M Eggo, Kevin Wing, Angel YS Wong, Harriet Forbes, John Tazare, John Parry, Frank Hester, Sam Harper, Ian J Douglas, Stephen JW Evans, Liam Smeeth, Ben Goldacre.

## Studies

Generation Scotland: David J Porteous, Drew Altschul, Chloe Fawns-Ritchie, Archie Campbell, Robin Flaig.

ALSPAC: Daniel J Smith.

Understanding Society: Michaela Benzeval.

TwinsUK: Deborah Hart, María Paz García, Rachel Horsfall

Centre for Longitudinal Studies: Matt Brown, Lisa Calderwood, Emla Fitzsimons, Alissa Goodman, Aida Sanchez

Born in Bradford: John Wright, Dan Mason

### Funding acknowledgements

This work was supported by the National Core Studies, an initiative funded by UKRI, NIHR and the Health and Safety Executive. The COVID-19 Longitudinal Health and Wellbeing National Core Study was funded by the Medical Research Council (MC_PC_20030).

The contributing studies have been made possible because of the tireless dedication, commitment and enthusiasm of the many people who have taken part. We would like to thank the participants and the numerous team members involved in the studies including interviewers, technicians, researchers, administrators, managers, health professionals and volunteers. We are additionally grateful to our funders for their financial input and support in making this research happen.

### Studies acknowledgements

Data gathered from questionnaire(s) was provided by Wellcome Longitudinal Population Study (LPS) COVID-19 Steering Group and Secretariat (221574/Z/20/Z).

Understanding Society is an initiative funded by the Economic and Social Research Council and various Government Departments, with scientific leadership by the Institute for Social and Economic Research, University of Essex, and survey delivery by NatCen Social Research and Kantar Public. The Understanding Society COVID-19 study is funded by the Economic and Social Research Council (ES/K005146/1) and the Health Foundation (2076161). The research data are distributed by the UK Data Service.

The Millennium Cohort Study, Next Steps, British Cohort Study 1970 and National Child Development Study 1958 are supported by the Centre for Longitudinal Studies, Resource Centre 2015-20 grant (ES/M001660/1) and a host of other co-funders. The COVID-19 data collections in these five cohorts were funded by the UKRI grant Understanding the economic, social and health impacts of COVID-19 using lifetime data: evidence from 5 nationally representative UK cohorts (ES/V012789/1).

The UK Medical Research Council and Wellcome (Grant Ref: 217065/Z/19/Z) and the University of Bristol provide core support for ALSPAC. A comprehensive list of grants funding is available on the ALSPAC website (http://www.bristol.ac.uk/alspac/external/documents/grant-acknowledgements.pdf). We are extremely grateful to all the families who took part in this study, the midwives for their help in recruiting them, and the whole ALSPAC team, which includes interviewers, computer and laboratory technicians, clerical workers, research scientists, volunteers, managers, receptionists and nurses. Please note that the study website contains details of all the data that is available through a fully searchable data dictionary and variable search tool” and reference the following webpage: http://www.bristol.ac.uk/alspac/researchers/our-data/. Ethical approval for the study was obtained from the ALSPAC Ethics and Law Committee and the Local Research Ethics Committees. Part of this data was collected using REDCap, see the REDCap website for details https://projectredcap.org/resources/citations/

TwinsUK receives funding from the Wellcome Trust (WT212904/Z/18/Z), the National Institute for Health Research (NIHR) Biomedical Research Centre based at Guy’s and St Thomas’ NHS Foundation Trust and King’s College London.The TwinsUK COVID-19 personal experience study was funded by the King’s Together Rapid COVID-19 Call award, under the projects original title ‘Keeping together through coronavirus: The physical and mental health implications of self-isolation due to the Covid-19 TwinsUK is also supported by the Chronic Disease Research Foundation and Zoe Global Ltd. The funders had no role in study design, data collection and analysis, decision to publish, or preparation of the manuscript.

Generation Scotland received core support from the Chief Scientist Office of the Scottish Government Health Directorates [CZD/16/6] and the Scottish Funding Council [HR03006]. Genotyping of the GS:SFHS samples was carried out by the Genetics Core Laboratory at the Wellcome Trust Clinical Research Facility, Edinburgh, Scotland and was funded by the Medical Research Council UK and the Wellcome Trust (Wellcome Trust Strategic Award “STratifying Resilience and Depression Longitudinally” (STRADL) Reference 104036/Z/14/Z). Generation Scotland is funded by the Wellcome Trust (216767/Z/19/Z) and (221574/Z/20/Z).

Born in Bradford (BiB) receives core infrastructure funding from the Wellcome Trust (WT101597MA), and a joint grant from the UK Medical Research Council (MRC) and UK Economic and Social Science Research Council (ESRC) (MR/N024397/1),the British Heart Foundation (BHF) (CS/16/4/32482), and The Health

Foundation COVID-19 award (2301201). The National Institute for Health Research Yorkshire and Humber Applied Research Collaboration (ARC) (NIHR200166), and Clinical Research Network both provide support for BiB research. Born in Bradford is only possible because of the enthusiasm and commitment of the children and parents in BiB. We are grateful to all the participants, health professionals, schools and researchers who have made Born in Bradford happen.

OpenSAFELY is jointly funded by UKRI, NIHR and Asthma UK-BLF [COV0076; MR/V015737/] and the Longitudinal Health and Wellbeing strand of the National Core Studies programme. EMIS and TPP provided technical expertise and infrastructure within their data environments pro bono in the context of a national emergency. The OpenSAFELY software platform is supported by a Wellcome Discretionary Award. BG’s work on clinical informatics is supported by the NIHR Oxford Biomedical Research Centre and the NIHR Applied Research Collaboration Oxford and Thames Valley. Funders had no role in the study design, collection, analysis, and interpretation of data; in the writing of the report; and in the decision to submit the article for publication. The views expressed are those of the authors and not necessarily those of the NIHR, NHS England, Public Health England or the Department of Health and Social Care.

### People funding

NJT is a Wellcome Trust Investigator (202802/Z/16/Z), is the PI of the Avon Longitudinal Study of Parents and Children (MRC & WT 217065/Z/19/Z), is supported by the University of Bristol NIHR Biomedical Research Centre, the MRC Integrative Epidemiology Unit (MC_UU_00011/1) and works within the CRUK Integrative Cancer Epidemiology Programme (C18281/A29019). SVK acknowledges funding from a NRS Senior Clinical Fellowship (SCAF/15/02), the Medical Research Council (MC_UU_00022/2) and the Scottish Government Chief Scientist Office (SPHSU17). ASFK acknowledges funding from the ESRC (ES/V011650/1). EJT acknowledges funding from the Wellcome Trust (WT212904/Z/18/Z). RM acknowledges support from the Elizabeth Blackwell Institute for Health Research, University of Bristol, and the Wellcome Trust Institutional Strategic Support Fund (204813/Z/16/Z). GBP acknowledges funding from the Economic and Social Research Council (ES/V012789/1). CLN acknowledges funding from the Medical Research Council (MR/R024774/1). KT works in a Unit that is supported by the University of Bristol and UK Medical Research Council (MC_UU_00011/3). DMW is supported by funding from UK Medical Research Council (MC_PC_20030). NC is supported by funding from the UK Medical Research Council (MC_UU_00019/2).

## List of Tables and Figures

**Table 1**. Descriptives of the ten LS analytic samples

**Table 2**. Counts and percentages of self-reported COVID-19 symptom length in the ten LS samples

**Table 3**. EHR table

**Figure 1**. Age plot

**Figure 2**. Risk factors for long COVID combining MA and OpenSAFELY

