## Supplementary Tables and Figures for "Risk factors for long COVID: analyses of 10 longitudinal studies and electronic health records in the UK"

**Risk factors for ongoing symptomatic COVID-19 and post-COVID syndrome in the population: analyses of 10 longitudinal studies and electronic health records in the UK**

Ellen J. Thompson†^*, Dylan M. Williams †^*, Alex J. Walker†^, Ruth E. Mitchell^, Claire L. Niedzwiedz^, Tiffany C. Yang^, Charlotte F. Huggins^, Alex S. F. Kwong^, Richard J. Silverwood, Giorgio Di Gessa, Ruth C. E. Bowyer, Kate Northstone, Bo Hou, Michael J. Green, Brian Dodgeon, Katie J. Doores, Emma L. Duncan, Frances M. K. Williams, OpenSAFELY Collaborative, Andrew Steptoe, David J. Porteous, Rosemary R. C. McEachan, Laurie Tomlinson, Ben Goldacre, Praveetha Patalay, George B. Ploubidis, Srinivasa Vittal Katikireddi, Kate Tilling, Christopher T. Rentsch, Nicholas J Timpson, Nishi Chaturvedi, Claire J. Steves*

† Joint first

^ Lead analyst team

### **List of Supplementary Tables**

**Supplementary Table S1***.* Details of each study

**Supplementary Table S2.** Ethics and data access statements for each study

**Supplementary Table S3**. Descriptives of analytic sample (self-reported COVID-19 symptoms) and non-analytic sample (no self-reported COVID-19 symptoms)

**Supplementary Table S4.** Length of time of symptoms (using self-report direct measures) by COVID-19 status (self-report)

**Supplementary Table S5.** Length of time of symptoms by COVID-19 status (swab/saliva and antibody test)

**Supplementary Table S6.** Length of time of symptoms (using sum of individual symptoms) by COVID-19 status (BIB; TwinsUK)

### **List of Supplementary Figures**

**Supplementary figure 1:** Categorical age associations with symptoms 4+ week in a sub-set of the longitudinal studies

**Supplementary figure 2:** Categorical age associations with symptoms 12+ week in a sub-set of the longitudinal studies

**Supplementary figure 3:** Full meta-analysis results for sociodemographic characteristics with symptoms for 4+ weeks in the longitudinal studies

**Supplementary figure 4:** Full meta-analysis results for health traits with symptoms for 4+ weeks in the longitudinal studies

**Supplementary figure 5:** Full meta-analysis results for sociodemographic characteristics with symptoms for 12+ weeks in the longitudinal studies

**Supplementary figure 6:** Full meta-analysis results for health traits with symptoms for 12+ weeks in the longitudinal studies

**Supplementary figure 7:** Secondary meta-analysis results for sociodemographic characteristics with symptoms for 4+ weeks in the longitudinal studies, including inverse probability weights for COVID-19 risk

**Supplementary figure 8:** Secondary meta-analysis results for health traits with symptoms for 4+ weeks in the longitudinal studies, including inverse probability weights for COVID-19 risk

**Supplementary figure 9:** Secondary meta-analysis results for sociodemographic characteristics with symptoms for 12+ weeks in the longitudinal studies, including inverse probability weights for COVID-19 risk

**Supplementary figure 10:** Secondary meta-analysis results for health traits with symptoms for 12+ weeks in the longitudinal studies, including inverse probability weights for COVID-19 risk

### **List of Supplementary Information**

**Supplementary information 1:** Information governance and ethics for the OpenSAFELY platform

**Supplementary Information 2:** Detail of method to derive long COVID by monthly symptom reporting

**Supplementary information 3:** Detail of method to derive inverse probability weights (IPW)

### **Supplementary Table S1**. Details of each study

| Study Population | Design and Sample Frame | 2020 Age Range | Pre-pandemic Survey | Details of Covid surveys  (response rate) | Analytic N |
| --- | --- | --- | --- | --- | --- |
| *Age Homogenous Cohorts* | |  |  |  |  |
| MCS: Millennium Cohort Study^1,2^ | Cohort of UK children born between Sept 2000 and Jan 2002 with regular follow-up surveys from birth. | 18-20 | 2018 | Spring 2021 survey response with the issued sample: 33.1% | 1055 |
| ALSPAC (G1): Avon Longitudinal Study of Parents and Children- Generation 1^3^ | Cohort of children born in the South-West of England between April 1991 and Dec 1992, with regular follow-up surveys from birth.  (original young people) | 27-29 | 2017-2018 | Three questionnaires: April (19%), June (17.4%), December (26.4%) | 668 |
| NS: Next Steps, formerly known as Longitudinal Study of Young People in England^1,4^ | Sample recruited via secondary schools in England at around age 13 with regular follow-up surveys thereafter. | 29-31 | 2015 | Spring 2021 survey response with the issued sample: 34.3% | 848 |
| BCS70: British Cohort Study 1970^1,5^ | Cohort of all children born in Great Britain (i.e. England, Wales & Scotland) in one week in 1970, with regular follow-up surveys from birth. | 50 | 2016 | Spring 2021 survey response with the issued sample: 45.4% | 889 |
| NCDS: National Child Development Study^1,6^ | Cohort of all children born in Great Britain (i.e. England, Wales & Scotland) in one week in 1958, with regular follow-up surveys from birth. | 62 | 2013 | Spring 2021 survey response with the issued sample: 58.5% | 709 |
| *Age Heterogeneous Studies* | |  |  |  |  |
| BIB: Born in Bradford^7,8^ | Birth cohort recruiting pregnant women and their children between 2007 and 2010 | 28-55 | 2016-2020 | Two surveys: April-Jun (30.7%) & Oct-Nov (39.9%) | 110 |
| USOC: Understanding Society: the UK Household Longitudinal Survey^9^ | A nationally representative longitudinal household panel study, based on a clustered-stratified probability sample of UK households, with all adults aged 16+ in chosen households surveyed annually. | 16-96 | 2018-2019 | Seven surveys (full/partial interview): April 2020 (42.0%); May (35.1%); Jun (33.5%); July (32.6%); Sep (30.6%), Nov (28.6%), Jan 2021 (28.5%) | 1033 |
| GS: Generation Scotland: the Scottish Family Health Study^10^ | A family-structured, population-based Scottish cohort, with participants aged 18-99 recruited between 2006-2011 | 27-100 | 2006-2011 | Two surveys: April-Jun (21.6%) & Jul-Aug (15.6%) | 343 |
| ALSPAC(G0): Avon Longitudinal Study of Parents and Children- Generation 0^11^ | Parents of the ALSPAC(G1) cohort described above, treated as a separate age-heterogenous study population.  (original parents) | 45-81 | 2011-2013 | Three questionnaires: April (12.4%), June (12.2%), December (14.3%) | 446 |
| TWINSUK: the UK Adult Twin Registry^12,13^ | A cohort of UK volunteer adult twins (55% monozygotic and 43% dizygotic) who were sampled between 18-101 years of age. | 22-96 | 2017-2018 | Three surveys: April (64.3%), July (77.6%) & November (76.1%) | 806 |

### **Supplementary Table S2.** Ethics and data access statements for each study

| NCDS, BCS70, NS and MCS | The most recent sweeps of the NCDS, BCS70, Next Steps and MCS have all been granted ethical approval by the National Health Service (NHS) Research Ethics Committee and all participants have given informed consent. Data for NCDS (SN 6137), BCS70 (SN 8547), Next Steps (SN 5545), MCS (SN 8682) and all four COVID-19 surveys (SN 8658) are available through the UK Data Service. NSHD data are available on request to the NSHD Data Sharing Committee. Interested researchers can apply to access the NSHD data via a standard application procedure. Data requests should be submitted to; further details can be found at <http://www.nshd.mrc.ac.uk/data.aspx>. doi:10.5522/NSHD/Q101; doi:10.5522/NSHD/Q10. |
| --- | --- |
| ALSPAC | Ethical approval was obtained from the ALSPAC Ethics and Law Committee and the Local Research Ethics Committees. The study website contains details of all the data that is available through a fully searchable data dictionary and variable search tool: <http://www.bristol.ac.uk/alspac/researchers/our-data>. ALSPAC data is available to researchers through an online proposal system. Information regarding access can be found on the ALSPAC website (<http://www.bristol.ac.uk/media-library/sites/alspac/documents/researchers/data-access/ALSPAC_Access_Policy.pdf>). |
| BIB | Ethical approval for Born in Bradford was granted by the National Health Service Health Research Authority Yorkshire and the Humber (Bradford Leeds) Research Ethics Committee (reference: 16/YH/0320). Data from the various BiB family studies are available to researchers; see the study website for information on how to access data (<https://borninbradford.nhs.uk/research/how-to-access-data/>). |
| USOC | The University of Essex Ethics Committee has approved all data collection for the Understanding Society main study and COVID-19 waves. No additional ethical approval was necessary for this secondary data analysis. All data are available through the UK Data Service (SN 6614 and SN 8644). |
| GS | Generation Scotland obtained ethical approval from the East of Scotland Committee on Medical Research Ethics (on behalf of the National Health Service). Reference number 20/ES/0021. Access to data is approved by the Generation Scotland Access Committee. See <https://www.ed.ac.uk/generation-scotland/for-researchers/access> or for further details. |
| TWINSUK | All wave of TwinsUK have received ethical approval associated with TwinsUK Biobank (19/NW/0187), TwinsUK (EC04/015) or Healthy Ageing Twin Study (H.A.T.S) (07/H0802/84) studies from NHS Research Ethics Committees at the Department of Twin Research and Genetic Epidemiology, King’s College London. The TwinsUK Resource Executive Committee (TREC) oversees management, data sharing and collaborations involving the TwinsUK registry (for further details see <https://twinsuk.ac.uk/resources-for-researchers/access-our-data/>). |

### **Supplementary Table S3**. Descriptives of analytic sample (self-reported COVID-19 symptoms) and non-analytic sample (no self-reported COVID-19 symptoms)

|  | **MCS** | | **ALSPAC G1** | | **Next Steps** | | **BIB** | | **Usoc** | | **TwinsUK** | | **GS** | | **ALSPAC G0** | | **BCS70** | | **NCDS** | |
| --- | --- | --- | --- | --- | --- | --- | --- | --- | --- | --- | --- | --- | --- | --- | --- | --- | --- | --- | --- | --- |
|  | Analysis sample | Excluded | Analysis sample | Excluded | Analysis sample | Excluded | Analysis sample | Excluded | Analysis sample | Excluded | Analysis sample | Excluded | Analysis sample | Excluded | Analysis sample | Excluded | Analysis sample | Excluded | Analysis sample | Excluded |
| Sample size | 1055 | 3,293 | 668 | 3446 | 848 | 3,317 | 110 | 488 | 1033 | 9267 | 806 | 4610 | 343 | 2805 | 446 | 3890 | 889 | 4,815 | 709 | 6,063 |
| Age, mean years (SD) | 19.9 (0.3) | 19.9 (0.3) | 28.4 | 28.4 | 31.0 (0.3) | 31.0 (0.3) | 40.7 (5.9) | 41.2 (5.7) | 48.5 (14.8) | 56.4 (16.1) | 52.7 (15.85) | 59.9 (15.78) | 56.0 (10.6) | 61.0 (11.5) | 58.3 (4.4) | 59.5 (4.8) | 51 * | 51 * | 63 * | 63 * |
| Female sex, % | 61.8 | 60.3 | 63.8 | 66.6 | 61.8 | 62.3 | 96.4 | 94.3 | 65.3 | 57.5 | 88 | 87.8 | 63.9 | 63.8 | 67.9 | 70.8 | 57 | 57.7 | 54.9 | 53.4 |
| **Ethnicity, %** |  |  |  |  |  |  |  |  |  |  |  |  |  |  |  |  |  |  |  |  |
| White | 81.7 | 83.3 | 95.5 | 95.8 | 67.7 | 73.4 | 44.5 | 47.3 | 85.1 | 89.9 | 96.2 | 97.1 | 96.2 | 98.3 | 98.4 | 98.0 | 84 | 85.5 | 92 | 91.2 |
| Non-white ethnic minority | 18.2 | 16.2 | 4.5 | 4.1 | 30 | 24.9 | 50.9 | 49.8 | 13.2 | 8.7 | 3.7 | 2.9 | 1.5 | 0.6 | 1.4 | 1.9) | 3 | 2.1 | 2.7 | 1.4 |
| Missing | 0.1 | 0.6 | 0 | 0.1 | 2.4 | 1.8 | 4.6 | 2.9 | 1.7 | 1.4 | 0.1 | 0.2 | 2.3 | 1.1 | 0.2 | 0.1 | 12.9 | 12.5 | 5.4 | 9.4 |
| **Education, %** |  |  |  |  |  |  |  |  |  |  |  |  |  |  |  |  |  |  |  |  |
| Degree | 46.8 | 39.3 | 50.6 | 51.4 | 49.7 | 43.3 | 10 | 9.6 | 48.4 | 39.5 | 49.9 | 45.5 | 49 | 45 | 23.8 | 26.6 | 42.4 | 42.3 | 40.1 | 41.3 |
| No degree | 47.6 | 54.8 | 22.3 | 24.2 | 42.2 | 46.7 | 74.5 | 76 | 41.5 | 46.7 | 27.8 | 41.1 | 49 | 53.1 | 68.8 | 65.5 | 49.9 | 50.9 | 58.5 | 56.4 |
| Missing | 5.6 | 5.9 | 27.1 | 24.4 | 11.1 | 10 | 15.5 | 14.3 | 10.1 | 13.8 | 22.3 | 13.4 | 2 | 1.9 | 7.4 | 7.8 | 7.7 | 6.9 | 1.4 | 2.4 |
| **IMD quintile, %** |  |  |  |  |  |  |  |  |  |  |  |  |  |  |  |  |  |  |  |  |
| 1 | 15.5 | 18.7 | 35 | 34.1 | 23.7 | 18.5 | 45.5 | 46.3 | - | - | 7.4 | 6.3 | 10.5 | 5.9 | 37.7 | 39.6 | 11.3 | 9.1 | 9.5 | 8.5 |
| 2 | 15.2 | 16.5 | 24.3 | 23.6 | 18.5 | 19.6 | 30 | 24.4 | - | - | 14.6 | 12.9 | 11.7 | 9.7 | 27.4 | 24.9 | 15.1 | 13.4 | 15.1 | 13.8 |
| 3 | 16.9 | 18.8 | 15.7 | 16.7 | 16.8 | 18.2 | 11.8 | 12.1 | - | - | 21.1 | 20.8 | 15.5 | 15.1 | 12.8 | 14.6 | 19.2 | 18.2 | 17.2 | 19.7 |
| 4 | 22 | 19.3 | 11.8 | 11.7 | 17.3 | 16.9 | 8.2 | 8.4 | - | - | 26.9 | 26.2 | 23 | 28.2 | 9.2 | 8.8 | 18.2 | 21.8 | 22.4 | 22.4 |
| 5 | 27.7 | 23.4 | 6.9 | 6.4 | 12.5 | 16.7 | 0.9 | 3.1 | - | - | 29.5 | 33 | 39.4 | 41 | 4.3 | 3.5 | 23.9 | 24.7 | 25.3 | 26.5 |
| Missing | 2.8 | 3.3 | 6.3 | 7.4 | 11.2 | 10.1 | 3.6 | 5.7 | - | - | 0.4 | 0.8 | -- | -- | 8.7 | 8.5 | 12.4 | 12.8 | 10.6 | 9.1 |
| **Social class, %** |  |  |  |  |  |  |  |  |  |  |  |  |  |  |  |  |  |  |  |  |
| Managerial, Admin, Professional | NA | NA | 18 | 14.9 | NA | NA | 23.6 | 28.3 | 38.9 | 31.1 | na | na | 53.1 | 47.2 | 12.8 | 11.5 | NA | NA | NA | NA |
| Intermediate | NA | NA | 41.9 | 39.2 | NA | NA | 32.7 | 27.7 | 16.6 | 16.4 | na | na | 17.8 | 492 | 29.2 | 35.8 | NA | NA | NA | NA |
| Manual/Routine | NA | NA | 25.6 | 31.6 | NA | NA | 19.1 | 16.8 | 21.3 | 17.4 | na | na | 11.4 | 8.1 | 42.6 | 38.1 | NA | NA | NA | NA |
| Not in employment | NA | NA | 0.3 | 0.5 | NA | NA |  |  | 20.5 | 33.6 | na | na |  | 0 | 1.1 | 0.7 | NA | NA | NA | NA |
| Missing | NA | NA | 14.2 | 13.8 | NA | NA | 24.5 | 27.3 | 2.7 | 1.5 | na | na | 17.8 | 27.1 | 14.4 | 14 | NA | NA | NA | NA |
| **Country, %** |  |  |  |  |  |  |  |  |  |  |  |  |  |  |  |  |  |  |  |  |
| England | 70.7 | 66.4 | 100 | 100 | 97.6 | 96.5 | 100 | 100 | 83.8 | 79.9 | 92.7 | 91.4 | 1.2 | 0.4 | 100 | 100 | 86.6 | 82.7 | 86.5 | 82.1 |
| Scotland | 8.8 | 13.3 |  |  | 0.6 | 0.6 |  |  | 6 | 9.5 | 3.2 | 4.5 | 98.8 | 99.5 |  |  | 6.4 | 8.5 | 6.4 | 8.8 |
| Wales | 12.9 | 11.1 |  |  | 1.1 | 0.7 |  |  | 6.7 | 6 | 3 | 3.1 |  |  |  |  | 5 | 5 | 5.4 | 5.2 |
| Northern Ireland | 7.1 | 8.6 |  |  | 0.1 | 0.1 |  |  | 3.5 | 4.6 | 0.1 | 0.1 |  | 0 |  |  | 0 | 0.2 | 0.3 | 0.1 |
| Missing | 0.5 | 0.6 |  |  | 0.6 | 2 |  |  | 0 | 0 | 1 | 0.8 |  |  |  |  | 2 | 0 | 1.6 | 3.8 |
| **Pre-pandemic mental health,** mean scale score (SD) | NA | NA | 6.5 (6.2) | 6.9 (6.4) | NA | NA | 4.3 (5.4) | 3.0 (3.8) | 12.6 (6.2) | 10.8 (5.2) | 8.01 (6.13) | 7.36 (5.97) | 16.2 (8.6) | 14.8 (7.5) | 7.2 (5.6) | 6.3 (5.3) | NA | NA | NA | NA |
| **Pre-pandemic mental health categories, %** |  |  |  |  |  |  |  |  |  |  |  |  |  |  |  |  |  |  |  |  |
| Yes | 15.4 | 15.8 | 14.7 | 17.9 | 22.2 | 22.9 | 10 | 5.7 | 28.1 | 15.8 | 3.7 | 51.5 | 12 | 8.7 | 13.9 | 11.3 | 15 | 14.3 | 11.7 | 11.6 |
| No | 79.2 | 77.6 | 55.7 | 54.4 | 65.1 | 65.4 | 67.3 | 73 | 69.3 | 82.6 | 40.9 | 3.3 | 72.6 | 71.3 | 65 | 68.8 | 16.7 | 68.4 | 78.8 | 78.3 |
| Missing | 5.4 | 6.6 | 29.63 | 27.7 | 12.7 | 11.7 | 22.7 | 21.3 | 2.6 | 1.6 | 55.3 | 42.5 | 15.5 | 20.0 | 21.1 | 19.9 | 16.3 | 17.3 | 9.5 | 10.1 |
| **Self-reported health, %** |  |  |  |  |  |  |  |  |  |  |  |  |  |  |  |  |  |  |  |  |
| Excellent | NA | NA | 16.6 | 14.7 | 23.9 | 21.9 | 5.5 | 5.7 | 8.9 | 9.8 | 15.5 | 13.8 |  |  | 16.6 | 18 | 16.2 | 16.4 | 11.4 | 13.9 |
| Very Good | NA | NA | 36.8 | 35.7 | 35.3 | 36.7 | 21.8 | 24.4 | 32.7 | 37.9 | 23.7 | 26.3 |  |  | 20.2 | 27.6 | 30.6 | 32.6 | 33.3 | 34.4 |
| Good | NA | NA | 19.6 | 23.8 | 20.8 | 21.7 | 30.9 | 35.9 | 32.5 | 32.6 | 16.00% | 18.4 |  |  | 30.9 | 27 | 26.1 | 24.2 | 29.1 | 29.2 |
| Fair | NA | NA | 4.3 | 5.1 | 60.5 | 6.9 | 13.6 | 10.2 | 15.7 | 13.7 | 4.8 | 5.9 |  |  | 7.4 | 5.5 | 10.9 | 10.6 | 12.7 | 11.5 |
| Poor | NA | NA | 1.3 | 1.5 | 1.3 | 1.8 | 5.5 | 2.5 | 6.1 | 3.9 | 1 | 0.8 |  |  | 2.9 | 1.4 | 3.8 | 3.7 | 3.8 | 3.7 |
| Missing | NA | NA | 21.3 | 19.1 | 12.3 | 11 | 22.7 | 21.3 | 4.1 | 2.1 | 39 | 34.8 |  |  | 22 | 20.6 | 12.4 | 12.5 | 9.7 | 7.4 |
| **Pre-pandemic BMI, mean kg/m2 (SD)** | 23.3 (4.7) | 23.2 (4.7) | 25.1 (5.3) | 24.8 (5.1) | 25.4 (5.8) | 25.2 (5.3) | 26.8 (5.3) | 26.0 (5.5) | - | - | 26.2 (4.91) | 26.7 (5.17) | 26.8 (5.5) | 26.6 (5.0) | 27.2 (4.9) | 26.6 (4.8) | 28.7 (5.9) | 28.2 (5.3) | 27.6 (5.0) | 27.1 (5.1) |
| **Pre-pandemic BMI categories, %** |  |  |  |  |  |  |  |  |  |  |  |  |  |  |  |  |  |  |  |  |
| >18.5 | 4.6 | 6.9 | 1.8 | 2.1 | 1.5 | 2.4 | 1 | 1.6 | - | - | 1.4 | 1.1 | 1.5 | 0.7 | 0.2 | 0.7 | 0 | 0.3 | 0.5 | 0.5 |
| 18.5 - 24.9999 | 63.5 | 59.6 | 41 | 39.0 | 46.9 | 47.9 | 31.8 | 40.4 | - | - | 19.7 | 26.1 | 32.4 | 34.1 | 2 | 26.5 | 22.8 | 24.3 | 26.8 | 31.9 |
| 25 – 29.9999 | 15.5 | 16.6 | 16.5 | 16.3 | 21.7 | 20.9 | 21.8 | 22.7 | - | - | 16.5 | 20.4 | 34.4 | 28.7 | 25.3 | 25 | 29.7 | 29.3 | 35.3 | 35.6 |
| 30+ | 8.7 | 8.4 | 10.3 | 8.9 | 13.6 | 14 | 20.9 | 16.4 | - | - | 8.9 | 10.8 | 16 | 16.4 | 13.9 | 12.8 | 27.3 | 25.7 | 22.7 | 19.4 |
| Missing | 7.7 | 8.6 | 30.4 | 33.7 | 16.3 | 14.8 | 24.5 | 18.9 | - | - | 53.5 | 41.6 | 17.3 | 20.2 | 38.6 | 35 | 20.1 | 20.4 | 14.7 | 12.6 |
| **Diabetes, %** |  |  |  |  |  |  |  |  |  |  |  |  |  |  |  |  |  |  |  |  |
| No | NA | NA | 67.8 | 66.5 | NA | NA | NA | NA | 93 | 92 | 65.1 | 64.9 | 82.5 | 78.6 | 76.9 | 77.2 | 84.5 | 84.6 | 83.4 | 87.7 |
| Yes | NA | NA | 0.3 | 0.4 | NA | NA | 5.5 | 3.9 | 7 | 8 | 1.7 | 3.1 | 2 | 1.5 | 1.1 | 2.3 | 3.2 | 2.9 | 6.8 | 4.9 |
| Missing | NA | NA | 31.9 | 33.1 | NA | NA | 94.5 | 96.1 | - | - | 33.1 | 31.9 | 15.5 | 19.9 | 22 | 20.5 | 12.4 | 12.5 | 9.9 | 7.5 |
| **Hypertension, %** |  |  |  |  |  |  |  |  |  |  |  |  |  |  |  |  |  |  |  |  |
| No | NA | NA | 67.2 | 65.6 | NA | NA | NA | NA | 79.7 | 74.1 | 60.9 | 57.6 | 77.6 | 69.4 | 65.5 | 70.3 | NA | NA | 69 | 73.6 |
| Yes | NA | NA | 0.8 | 1.2 | NA | NA | 12.7 | 5.9 | 20.3 | 25.9 | 12.5 | 19.7 | 7 | 10.7 | 15.3 | 13.1 | NA | NA | 21.3 | 19.9 |
| Missing | NA | NA | 32.0 | 33.2 | NA | NA | 87.3 | 94.1 | - | - | 26.6 | 22.7 | 15.5 | 19.9 | 19.3 | 16.6 | NA | NA | 9.7 | 7.5 |
| **High cholesterol, %** |  |  |  |  |  |  |  |  |  |  |  |  |  |  |  |  |  |  |  |  |
| No | NA | NA | 67.1 | 63.5 | NA | NA | Na | NA | - | - | 58.4 | 57.6 |  |  | 52 | 54.9 | NA | NA | NA | NA |
| Yes | NA | NA | 0 | 0.1 | NA | NA | 1.8 | 1.8 | - | - | 15.1 | 23.2 |  |  | 6.5 | 7.6 | NA | NA | NA | NA |
| Missing | NA | NA | 32.9 | 36.4 | NA | NA | 98.2 | 98.2 | - | - | 26.4 | 22.2 |  |  | 41.5 | 37.5 | NA | NA | NA | NA |
| **Asthma, %** |  |  |  |  |  |  |  |  |  |  |  |  |  |  |  |  |  |  |  |  |
| No | 74.7 | 76.2 | 49.4 | 49.7 | NA | NA | NA | NA | 78.9 | 85.3 | 7.3 | 10.6 | 71.1 | 72 | 65.7 | 68 | 78 | 77.3 | 80.5 | 81.8 |
| Yes | 10.9 | 10.2 | 18 | 17.1 | NA | NA | 10.9 | 12.3 | 21.1 | 14.7 | 7.2 | 7.9 | 13.4 | 8.1 | 11.7 | 11.2 | 9.7 | 10.2 | 9.6 | 10.7 |
| Missing | 14.4 | 13.6 | 32.6 | 33.2 | NA | NA | 89.1 | 87.7 | - | - | 85.5 | 81.6 | 15.5 | 19.9 | 22.7 | 20.8 | 12.4 | 12.5 | 9.9 | 7.5 |
| **Current smoker, %** |  |  |  |  |  |  |  |  |  |  |  |  |  |  |  |  |  |  |  |  |
| No | 43.2 | 48.7 | 49.4 | 49.7 | 64.4 | 61.2 | 84.5 | 83.2 | 91.5 | 92.2 | 85.9 | 88.5 | 93.6 | 94.7 | 57.2 | 61.1 | 72.4 | 69 | 79 | 77.8 |
| Yes | 11.8 | 8.4 | 18 | 17.1 | 8 | 10.1 | 5.5 | 6.6 | 8.4 | 7.8 | 13.8 | 10.7 | 5.5 | 4.8 | 28.3 | 26.2 | 7.3 | 10 | 4.5 | 7.5 |
| Missing | 45 | 42.9 | 32.63 | 33.2 | 27.6 | 28.7 | 10 | 10.2 | 0.1 | 0.0 | 0.4 | 0.8 | 0.1 | 0.6 | 14.6 | 12.7 | 20.3 | 21 | 16.5 | 14.7 |

Sources: MCS (Millennium Cohort Study); ALSPAC G1 (Children of the Avon Longitudinal Study of Parents and Children); NS (Next Steps); BCS 70 (1970 British Cohort Study), NCDS (National Child Development Study); USoc (Understanding Society); GS (Generation Scotland: the Scottish Family Health Study); TwinsUK (UK Adult Twin Registry); ALSPAC G0 (parents of ALSPAC); BiB (Born in Bradford). Unweighted data. Note. * SD values for age are approximately zero for these cohorts. All participants in NCDS and BCS70 were born in the same week in 1958 and 1970, respectively

### **Supplementary Table S4.** Length of time of symptoms (using self-report direct measures) by COVID-19 status (self-report)

|  | Mean age | COVID-19 ascertainment | N with symptom duration data | Duration of symptoms, N (%) |  |  |
| --- | --- | --- | --- | --- | --- | --- |
|  |  |  |  | Acute (0-4 weeks) | Ongoing symptomatic COVID-19 (4-12 weeks) | Post COVID-19 syndrome (12+ weeks) |
| **MCS** | 19.9 | ***Confirmed*** | ***552*** | ***532 (96.4)*** | ***15 (2.7)*** | ***5 (0.9)*** |
|  |  | Suspected | 503 | 478 (95.0) | 17 (3.4) | 8 (1.6) |
| **ALSPAC G1** | 28.4 | *Confirmed* | *187* | *144 (77.0)* | *25 (13.4)* | *18 (9.6)* |
|  |  | Suspected | 481 | 375 (78.0) | 15 (22.0) | 34 (7.1) |
| **Next Steps** | 31 | *Confirmed* | *400* | *360 (90.0)* | *26 (6.5)* | *14 (3.5)* |
|  |  | Suspected | 448 | 413 (92.2) | 25 (5.6) | 10 (2.2) |
| **USoc** | 48.5 | *Confirmed* | *95* | *73 (76.8)* | *17 (17.9)* | *5 (5.3)* |
|  |  | Suspected | 351 | 229 (65.2) | 51 (17.1) | 71 (20.2) |
| **BCS70** | 51 | *Confirmed* | *386* | *316 (81.9)* | *50 (13.0)* | *20 (5.2)* |
|  |  | Suspected | 503 | 441 (87.7) | 34 (6.8) | 28 (5.6) |
| **TwinsUK** | 52.7 | *Confirmed* | *252* | *164 (65.1)* | *42 (16.7)* | *46 (18.3)* |
|  |  | Suspected | 550 | 412 (74.9) | 103 (18.7) | 35 (6.4) |
| **GS** | 56 | *Confirmed* | *83* | *51 (61.5)* | *19 (22.9)* | *13 (15.7)* |
|  |  | Suspected | 252 | 173 (67.7) | 35 (13.9) | 44 (17.5) |
| **ALSPAC G0** | 58.3 | *Confirmed* | *35* | *73 (76.8)* | *17 (17.9)* | *5 (5.3)* |
|  |  | Suspected | 77 | 229 (65.2) | 51 (14.5) | 71 (20.2) |
| **NCDS** | 63 | *Confirmed* | *313* | *248 (79.2)* | *49 (15.7)* | *16 (5.1)* |
|  |  | Suspected | 396 | 330 (83.3) | 48 (12.1) | 18 (4.6) |
| **BiB** | 41.2 | *Confirmed* | *34* | *22 (64.7)* | *8 (23.5)* | *4 (11.8)* |
|  |  | Suspected | 76 | 18 (23.7) | 17 (22.4) | 41 (53.9) |

Note. *Confirmed*: tested positive by PCR, antigen and/or antibody test; Suspected: strong personal suspicion and/or medical advice of COVID-19, but no test result to confirm

### **Supplementary Table S5.** Length of time of symptoms by COVID-19 status (swab/saliva and antibody test)

|  | COVID-19 ascertainment | N with symptom duration data | Duration of symptoms, N (%) |  |  |
| --- | --- | --- | --- | --- | --- |
|  |  |  | Acute (0-4 weeks) | Ongoing symptomatic COVID-19 (4-12 weeks) | Post COVID-19 syndrome (12+ weeks) |
| **ALSPAC G1** | Symptomatic test positive (antibody or PCR) and self-diagnosed | *193 (100)* | *150 (78)* | *21 (11)* | *22 (11)* |
|  | Symptomatic test negative (antibody or PCR) and self-diagnosed | 282 (100) | 215 (78) | 45 (16) | 22 (8) |
| **TwinsUK** | Symptomatic test positive (antibody or PCR) and self-diagnosed | *260(100)* | *160 (61.5)* | *50 (19.2)* | *50 (19.2)* |
|  | Symptomatic test negative (antibody or PCR) and self-diagnosed | 469 (100) | 360 (76.8) | 83 (17.7) | 26 (5.5) |
| **ALSPAC G0** | Symptomatic test positive (antibody or PCR) and self-diagnosed | *57(100)* | *44 (77.2)* | *5 (8.8)* | *8 (14)* |
|  | Symptomatic test negative (antibody or PCR) and self-diagnosed | 202 (100) | 131 (64.9) | 39 (19.3) | 32 (15.8) |
| **BiB** | Symptomatic test positive (antibody or PCR) and self-diagnosed | *35 (100)* | *23 (65.7)* | *8 (22.9)* | *4 (11.4)* |
|  | Symptomatic test negative (antibody or PCR) and self-diagnosed | 42 (100) | 6 (14.3) | 9 (21.4) | 27 (64.3) |

### **Supplementary Table S6.** Length of time of symptoms (using sum of individual symptoms) by COVID-19 status (BIB; TwinsUK)

|  | COVID-19 ascertainment | N with symptom duration data | Duration of symptoms, N (%) |  |  |
| --- | --- | --- | --- | --- | --- |
|  |  |  | Acute (0-4 weeks) | Ongoing symptomatic COVID-19 (4-12 weeks) | Post COVID-19 syndrome (12+ weeks) |
| **TwinsUK** | Not Confirmed | 4611 | 820 (17.8) | 1006 (21.8) | 1328 (28.8) |
|  | *Confirmed* | *363* | *87 (24)* | *104 (28.7)* | *139 (38.3)* |
|  | Suspected | 696 | 185 (26.6) | 142 (20.4) | 296 (42.5) |
| **BiB** | *Confirmed* | *34* | *22 (64.7)* | *8 (23.5)* | *4 (11.8)* |
|  | Suspected | 76 | 18 (23.7) | 17 (22.4) | 41 (53.9) |

**Supplementary figure 1:** Categorical age associations with symptoms 4+ week in a sub-set of the longitudinal studies and EHRs from OpenSAFELY

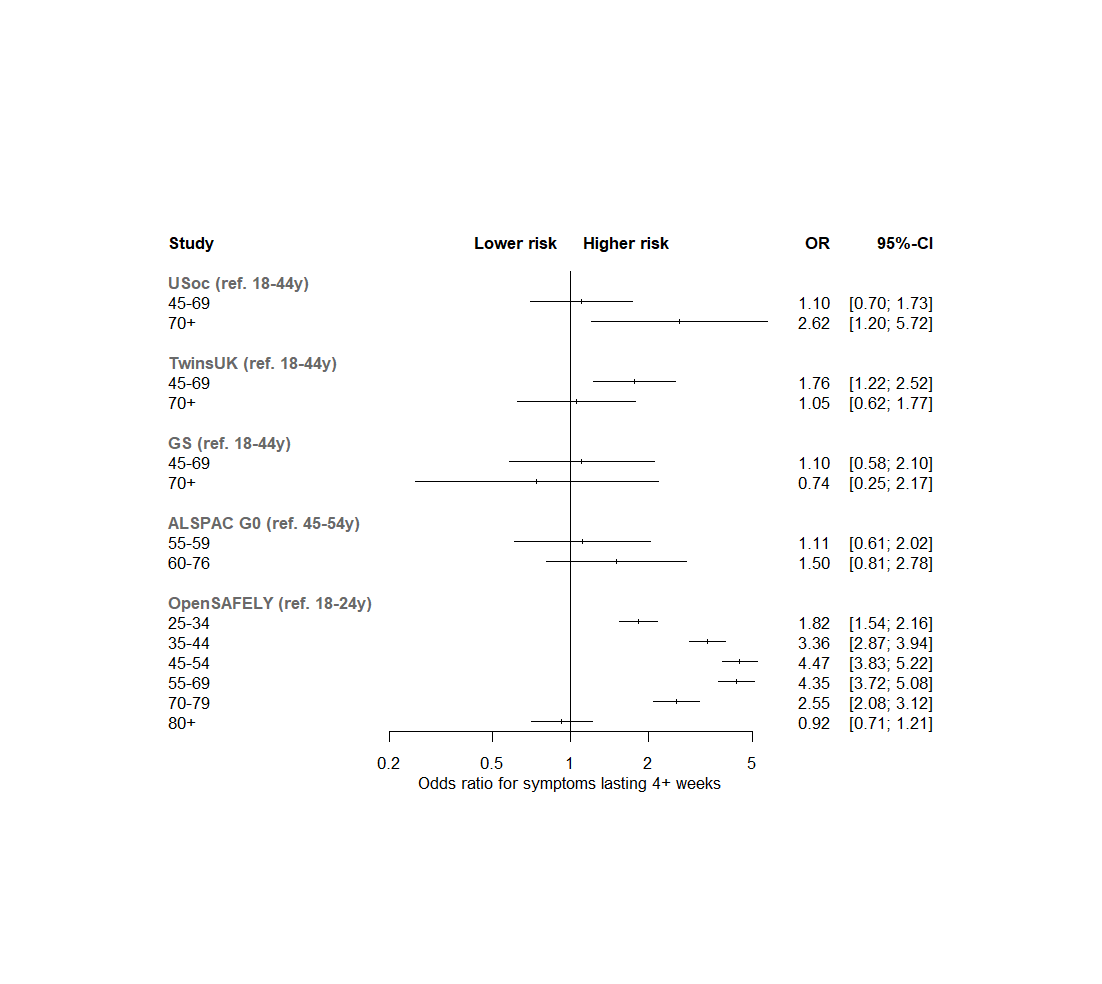

**Supplementary figure 2:** Categorical age associations with symptoms 12+ week in a sub-set of the longitudinal studies

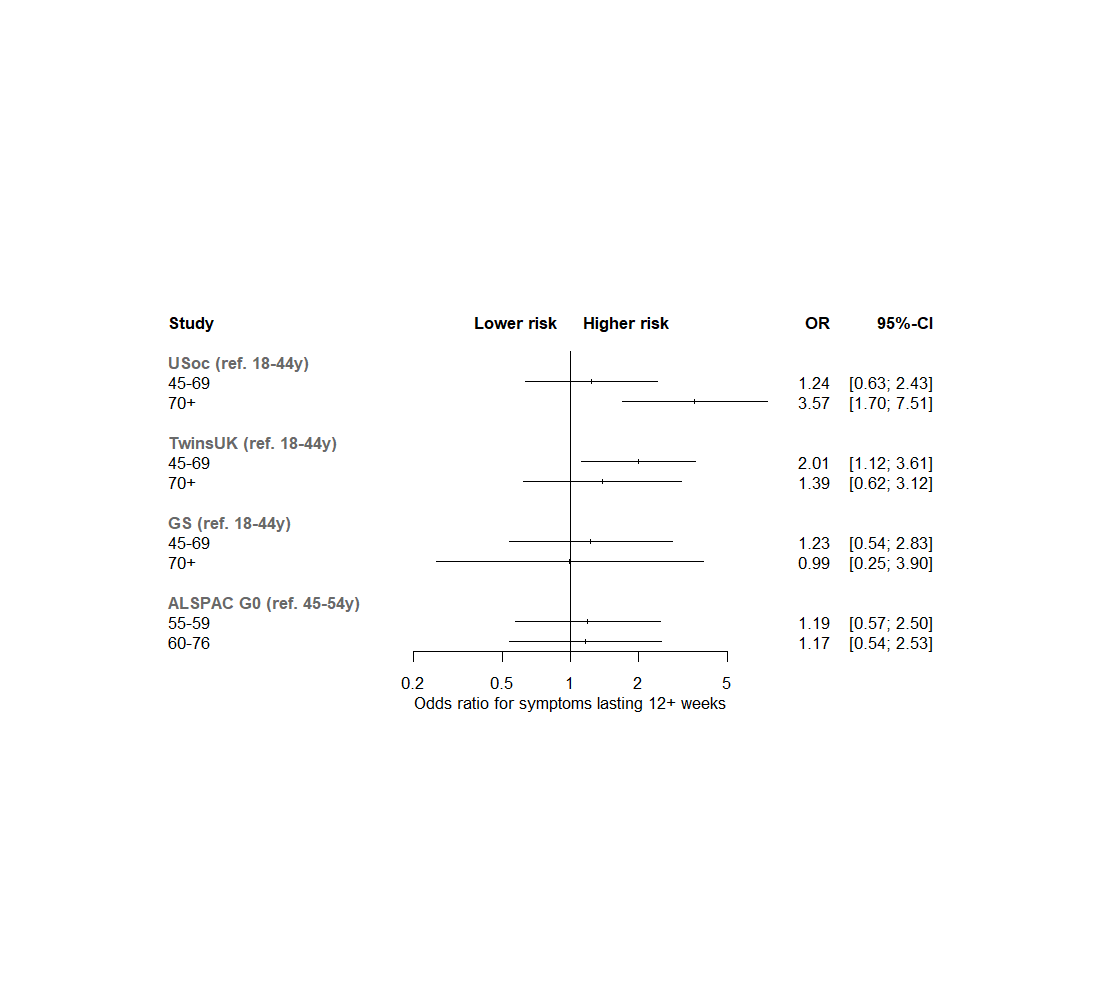

**Supplementary figure 3:** Full meta-analysis results for sociodemographic characteristics with symptoms for 4+ weeks in the longitudinal studies

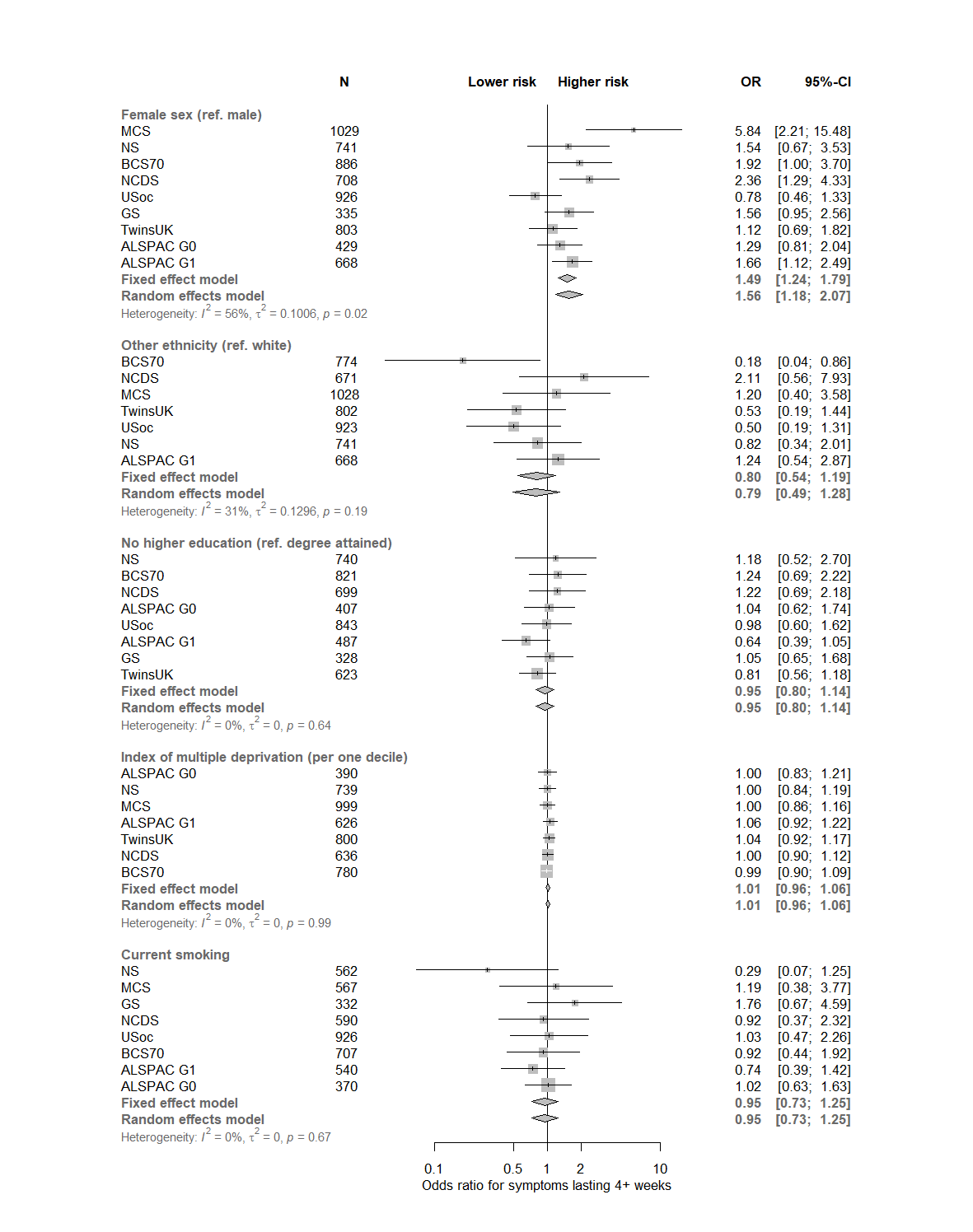

Associations adjusted for age and sex, where relevant

**Supplementary figure 4:** Full meta-analysis results for health factors with symptoms for 4+ weeks in the longitudinal studies

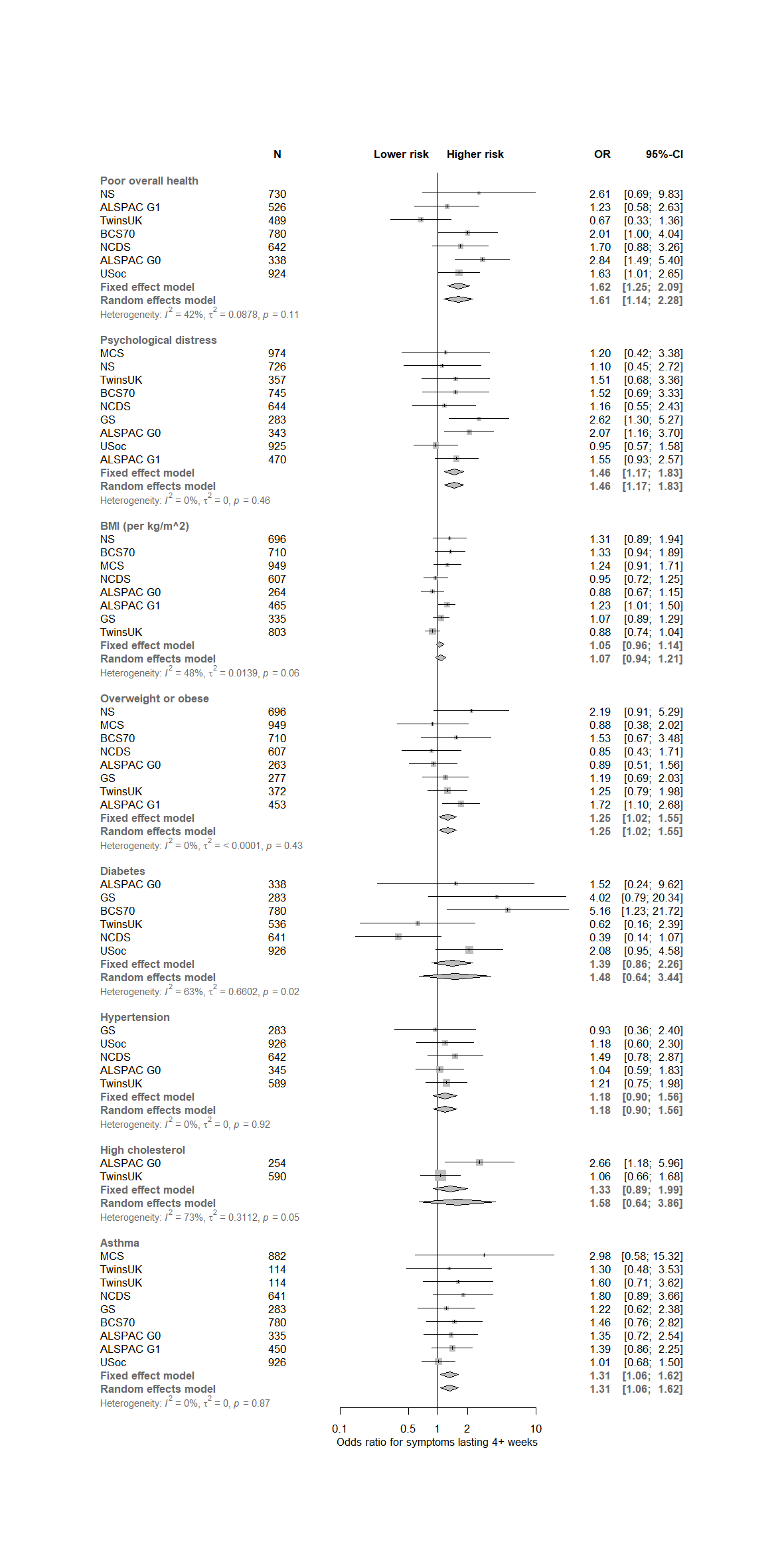

Associations adjusted for age and sex

**Supplementary figure 5:** Full meta-analysis results for sociodemographic characteristics with symptoms for 12+ weeks in the longitudinal studies

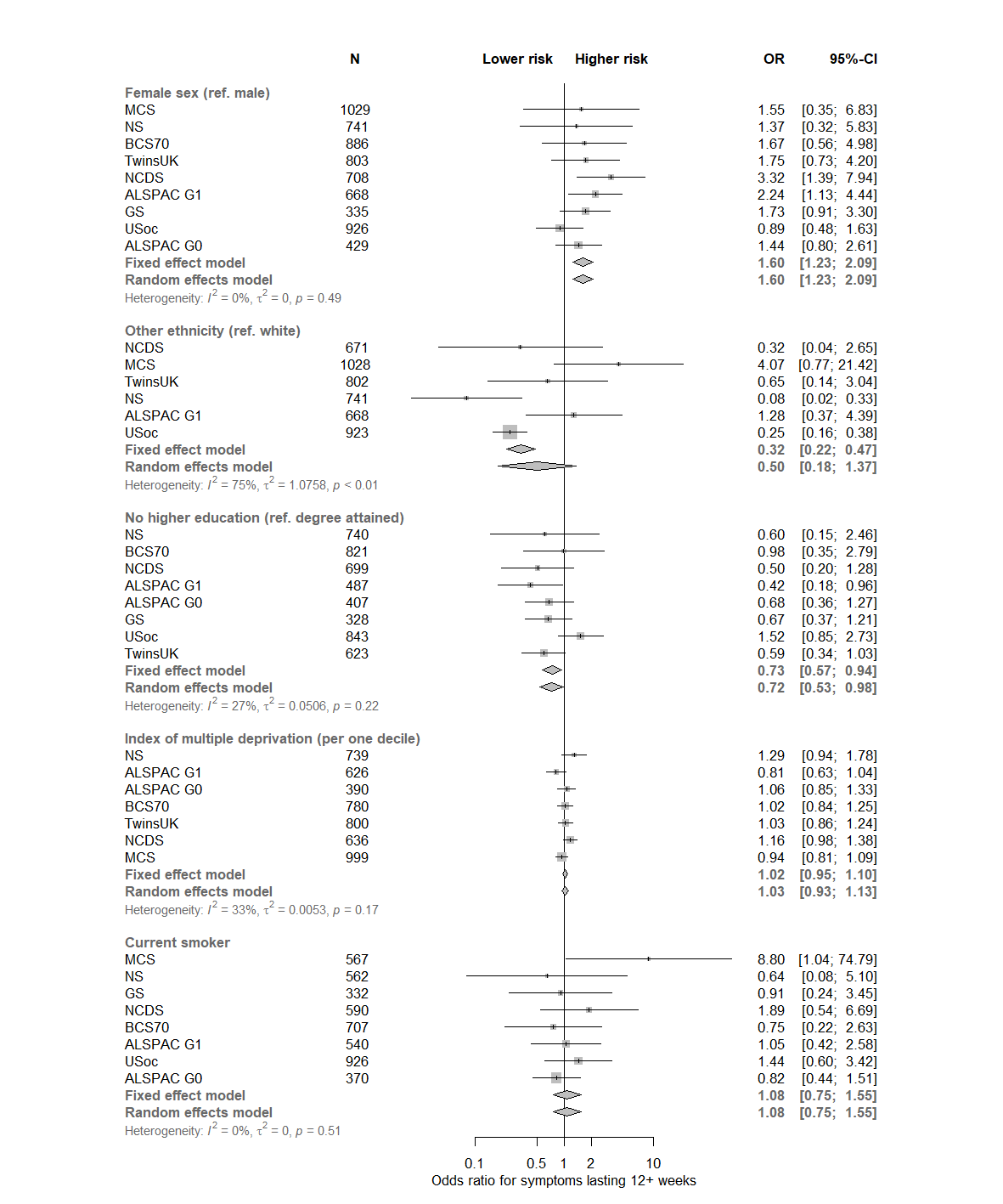

Associations adjusted for age and sex, where relevant

**Supplementary figure 6:** Full meta-analysis results for health factors with symptoms for 12+ weeks in the longitudinal studies

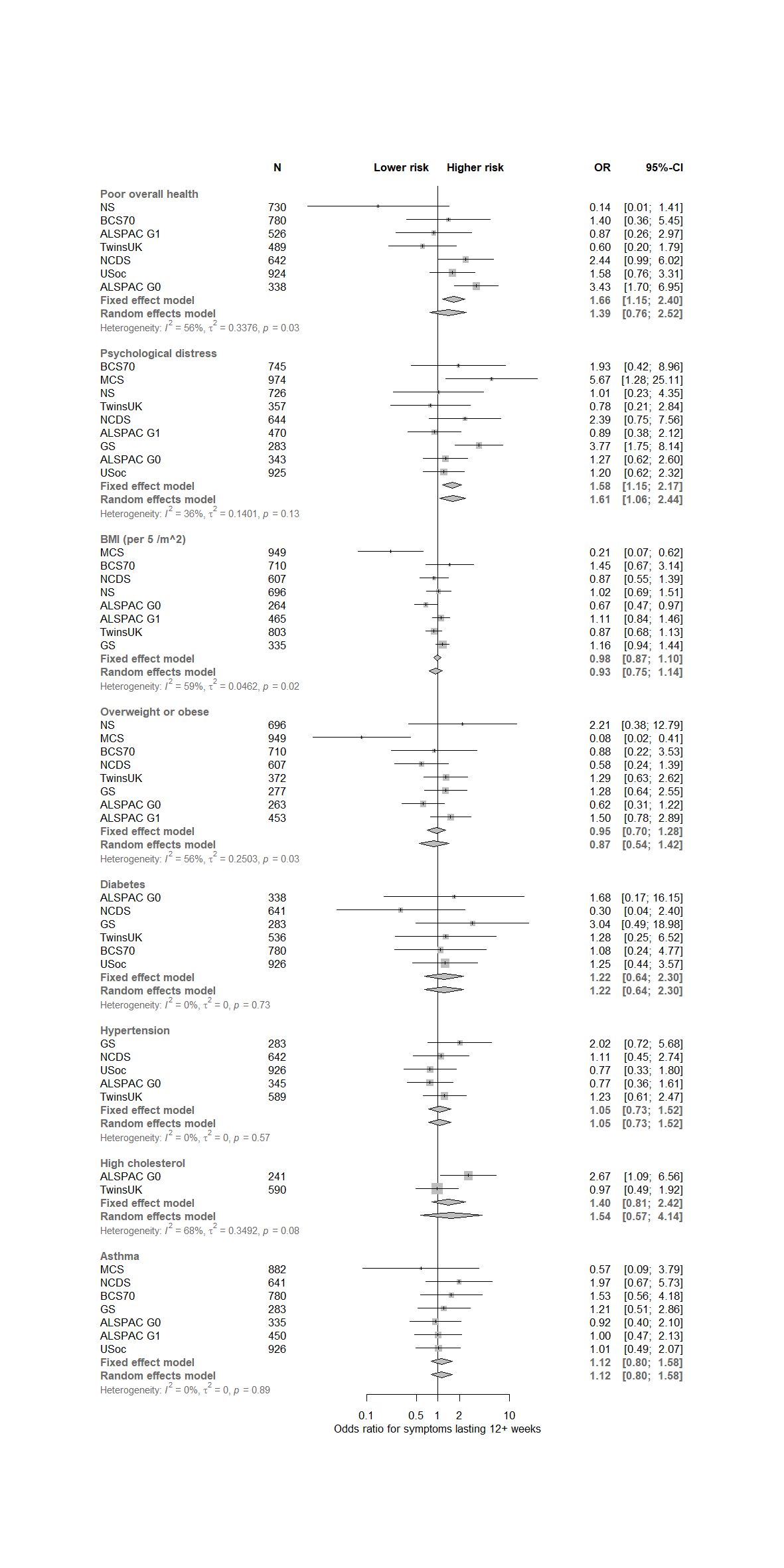

Associations adjusted for age and sex

**Supplementary figure 7:** Secondary meta-analysis results for sociodemographic characteristics with symptoms for 4+ weeks in the longitudinal studies, including inverse probability weights for COVID-19 risk

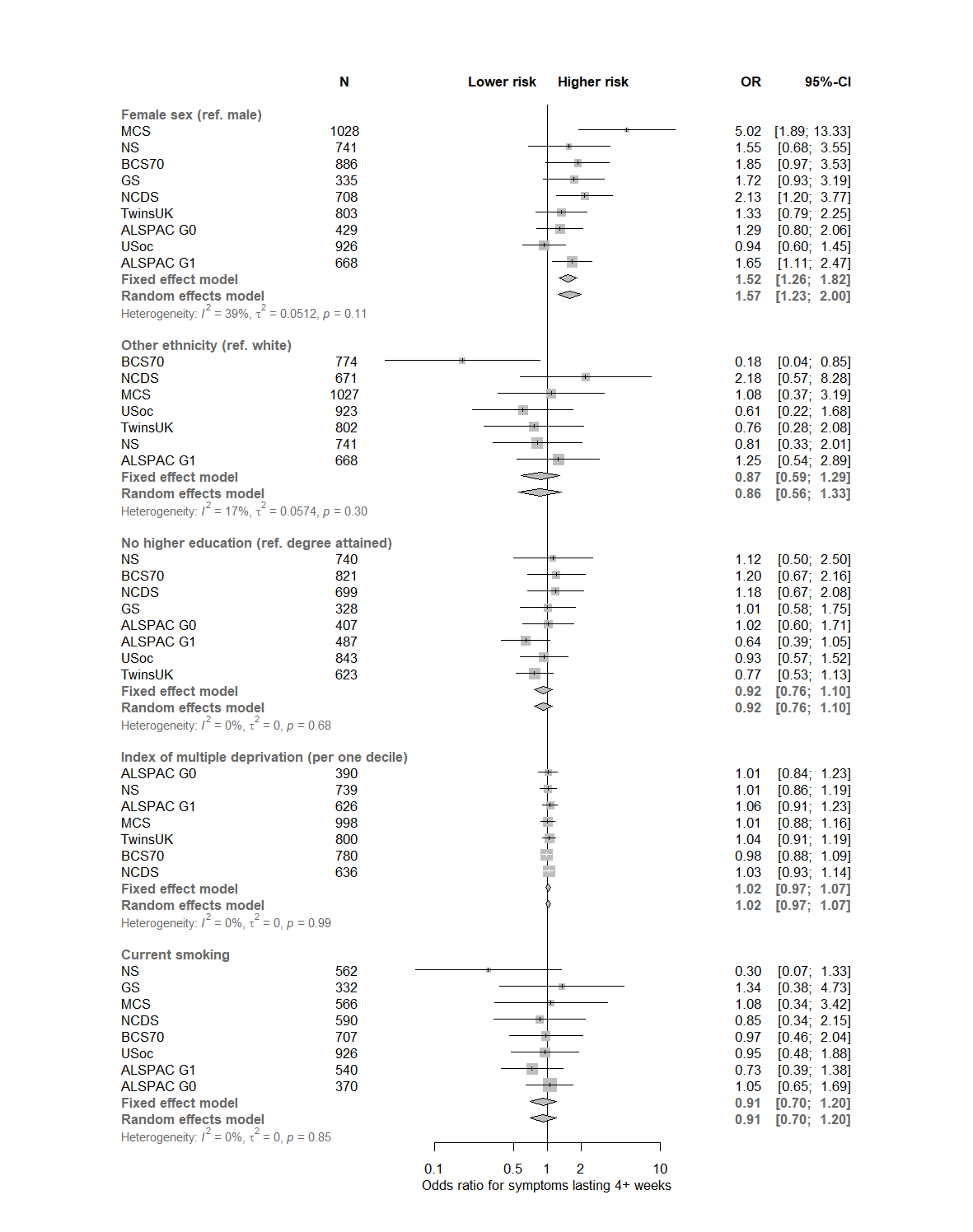

Associations adjusted for age and sex

**Supplementary figure 8:** Secondary meta-analysis results for health factors with symptoms for 4+ weeks in the longitudinal studies, including inverse probability weights for COVID-19 risk

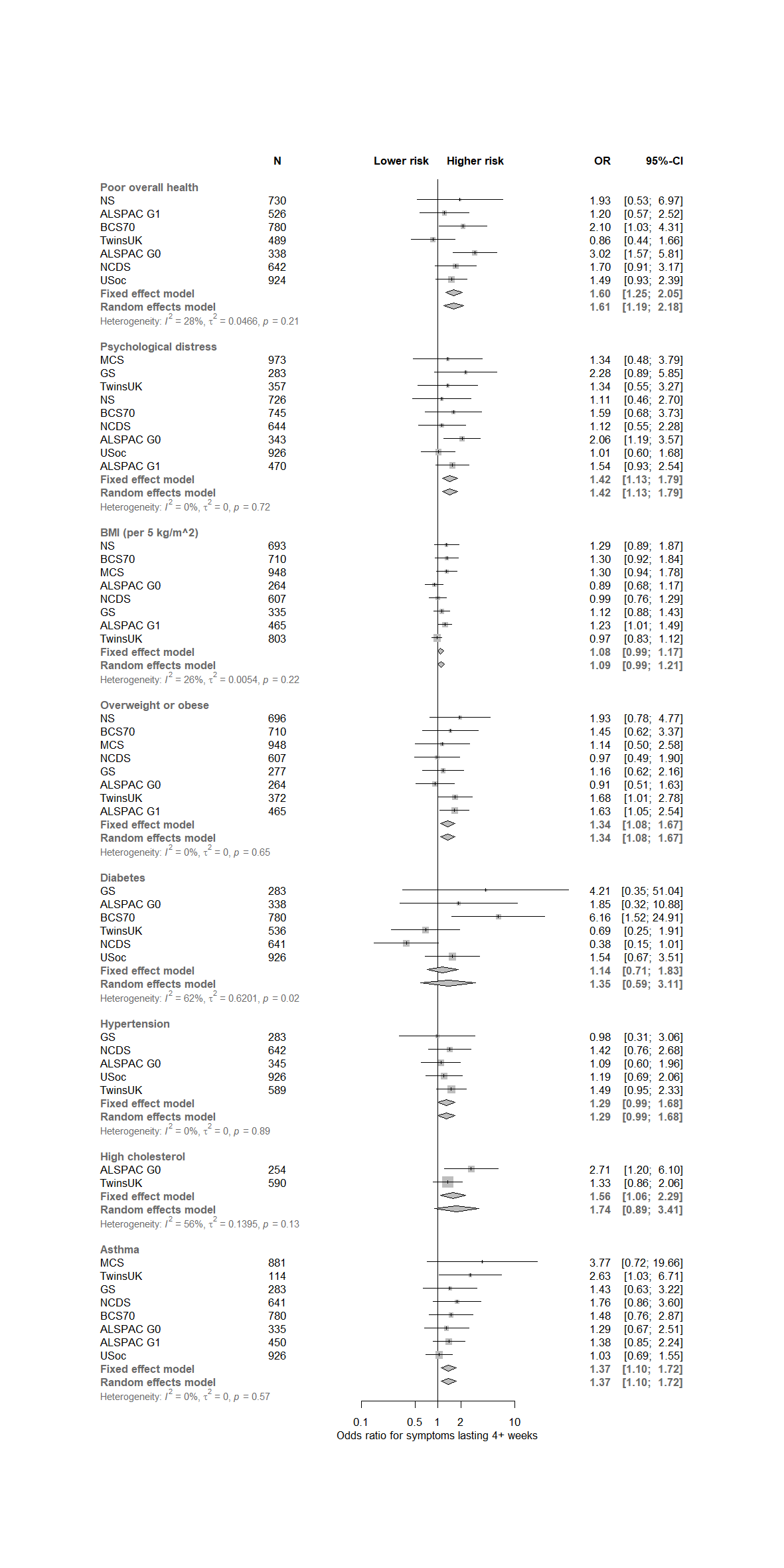

Associations adjusted for age and sex

**Supplementary figure 9:** Secondary meta-analysis results for sociodemographic characteristics with symptoms for 12+ weeks in the longitudinal studies, including inverse probability weights for COVID-19 risk

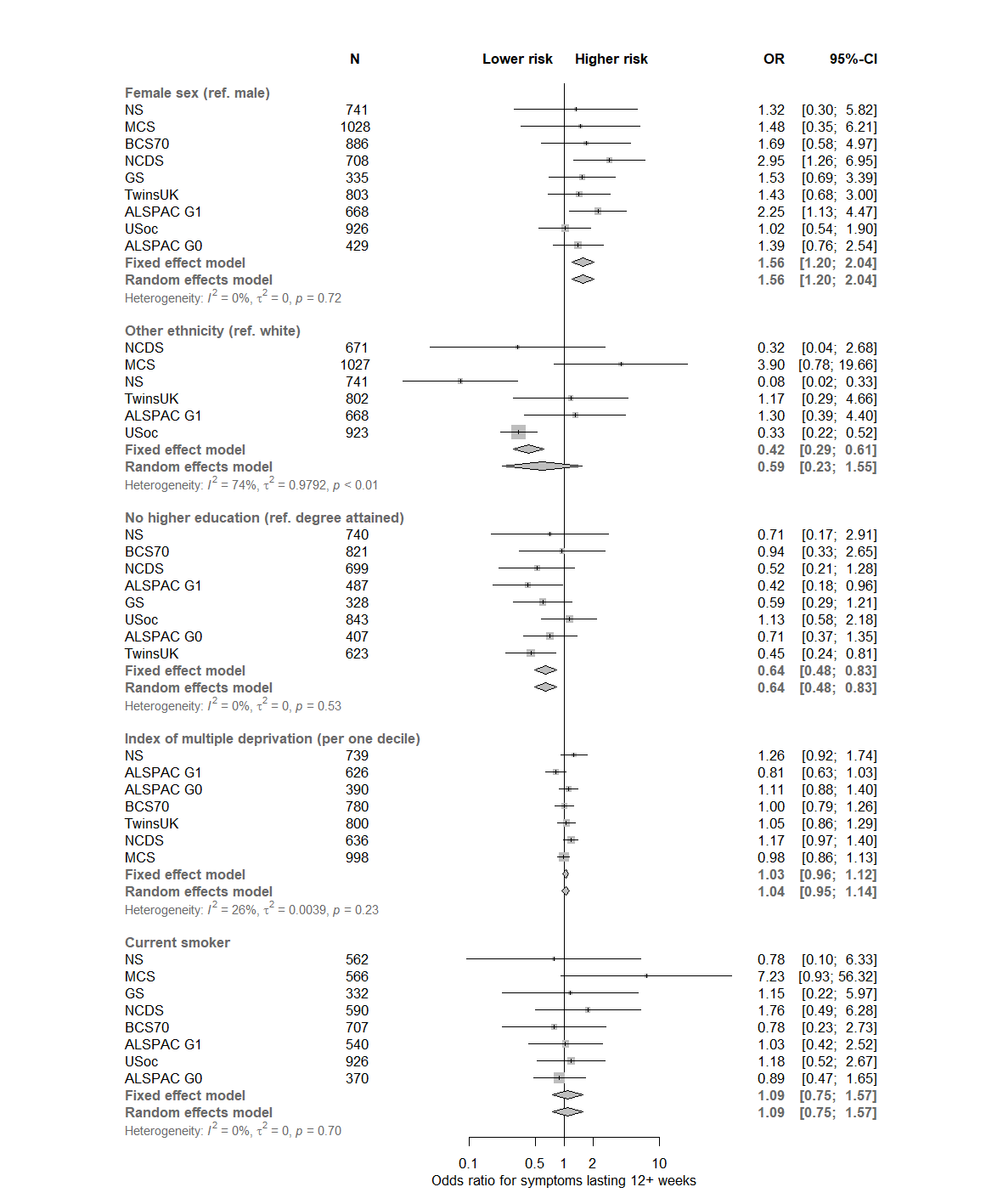

Associations adjusted for age and sex

**Supplementary figure 10:** Secondary meta-analysis results for health traits with symptoms for 12+ weeks in the longitudinal studies, including inverse probability weights for COVID-19 risk

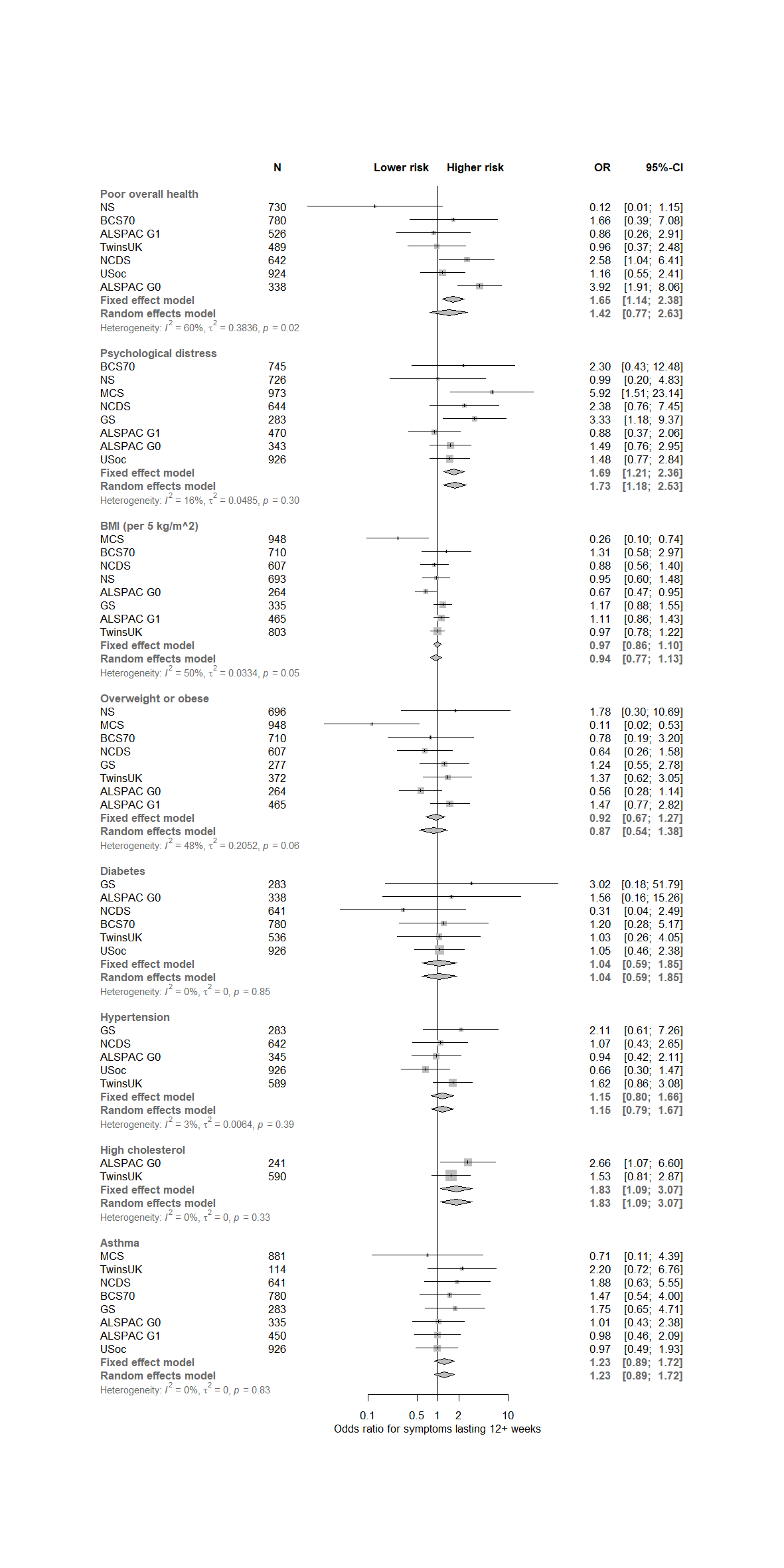

Associations adjusted for age and sex

**Supplementary information 1:** Information governance and ethics for the OpenSAFELY platform

NHS England is the data controller; TPP is the data processor; and the key researchers on OpenSAFELY are acting on behalf of NHS England. OpenSAFELY is hosted within the TPP environment which is accredited to the ISO 27001 information security standard and is NHS IG Toolkit compliant;1,2 patient data are pseudonymised for analysis and linkage using industry standard cryptographic hashing techniques; all pseudonymised datasets transmitted for linkage onto OpenSAFELY are encrypted; access to the platform is via a virtual private network (VPN) connection, restricted to a small group of researchers who hold contracts with NHS England and only access the platform to initiate database queries and statistical models. Pseudonymised structured data include demographics, medications prescribed from primary care, diagnoses, and laboratory measures. No free text data are included. All database activity is logged; only aggregate statistical outputs leave the platform environment following best practice for anonymisation of results such as statistical disclosure control for low cell counts.3 The OpenSAFELY research platform adheres to the obligations of the UK General Data Protection Regulation (GDPR) and the Data Protection Act 2018. In March 2020, the Secretary of State for Health and Social Care used powers under the UK Health Service (Control of Patient Information) Regulations 2002 (COPI) to require organisations to process confidential patient information for the purposes of protecting public health, providing healthcare services to the public and monitoring and managing the COVID-19 outbreak and incidents of exposure; this sets aside the requirement for patient consent.4 Taken together, these provide the legal bases to link patient datasets on the OpenSAFELY platform. GP practices, from which the primary care data are obtained, are required to share relevant health information to support the public health response to the pandemic and have been informed of the OpenSAFELY analytics platform. This study was approved by the Health Research Authority (REC reference 20/LO/0651) and by the LSHTM Ethics Board (ref 21863).

1	NHS Digital. Data Security and Protection Toolkit. 2020. https://digital.nhs.uk/data-and-information/looking-after-information/data-security-and-information-governance/data-security-and-protection-toolkit (accessed Aug 11, 2020).

2	NHS Digital. BETA - Data Security Standards. 2020. https://digital.nhs.uk/about-nhs-digital/our-work/nhs-digital-data-and-technology-standards/framework/beta---data-security-standards (accessed Aug 11, 2020).

3	NHS Digital. ISB1523: Anonymisation Standard for Publishing Health and Social Care Data. 2020. https://digital.nhs.uk/data-and-information/information-standards/information-standards-and-data-collections-including-extractions/publications-and-notifications/standards-and-collections/isb1523-anonymisation-standard-for-publishing-health-and-social-care-data (accessed Aug 11, 2020).

4	Secretary of State for Health-UK Government. Coronavirus (COVID-19): notification to organisations to share information. 2020. https://www.gov.uk/government/publications/coronavirus-covid-19-notification-of-data-controllers-to-share-information (accessed Aug 11, 2020).

**Supplementary Information 2:** Detail of method to derive long COVID by monthly symptom reporting

***BiB***

BiB study members who self-reported COVID-19 were asked to report whether any particular symptoms were present during March-September 2020. Specifically, participants were presented with 27 symptoms (i.e. “decrease in appetite”, “nausea and/or vomiting”, “diarrhoea”, “abdominal pain/tummy ache”, “sore eyes”, “loss of sense of smell or taste”, “sore throat”, “hoarse voice”, “headache”, “dizziness”, “new persistent cough”, “tightness in the chest”, “chest pain”, “shortness of breath”, “fever”, “chills”, “difficulty sleeping”, “felt more tired than normal”, “severe fatigue”, “numbness or tingling somewhere in the body”, “feeling of heaviness in arms or legs”, “achy muscles”, “raised, red, itchy areas on the skin”, “sudden swelling of the face or lips”) and asked to tick whether the symptoms were present for the months March – September 2020. Only 24 of the 27 symptoms were included for consideration as three symptoms (“runny nose”, “sneezing”, and “blocked nose”) were considered non-specific.

They were also asked: (1) whether they had symptoms in the past week (yes; no); (2) whether they think they have had COVID-19 (Yes, confirmed by a positive test; Yes, suspected by a doctor but not tested; Yes, my own suspicions; No) which was re-categorized to a binary response of “yes” if they responded affirmatively and “no” if they did not; (3) if yes to (2), when were you told / when did you think you had COVID-19 (write-in ‘reported positive’ date), and (4) whether they had a positive result from a swab (polymerase chain reaction [PCR]) test (yes; no; don’t know) or an antibody test (yes; no; don’t know) which was re-categorized to a binary variable with “yes” indicating a positive result from a swab or antibody test, “no” indicating a negative result from a swab or antibody test, and those responding “don’t know” coded as missing.

Data were used to derive symptom length categories above by summing included symptoms present for 0-4 weeks; 4-12 weeks or 12+ weeks. Participants were coded to the 0–4-week category if they selected symptoms in the same month as the reported positive date or the participant reported symptoms in the past week and it falls within the same month as positive date (for example, when the reported positive date is after the last symptom month of September and they reported symptoms in the past week).

Participants were coded to the 4–12-week category if they selected 2-3 symptom months. A 3-month duration was allowed only if one of the months is within the same month as the reported positive date.

Participants were coded to the 12+ week category if they selected 3+ symptom months. A 3-month duration was allowed only if the reported positive date was not one of the selected months.

The final analytical sample of long COVID participants (n=110) included only those who responded affirmatively to have had COVID-19 whether confirmed by a positive test, was suspected by doctor but not tested, or from their own judgment.

***TwinsUK***

In TwinsUK, all study members were asked to report whether they had experienced particular symptoms (33 in total) between February and November 2020. Specifically, twins were presented with 33 symptoms (i.e. “cold or flu symptoms”, “decrease in appetite”, “nausea and/or vomiting”, “diarrhoea”, “abdominal pain/stomach ache”, “runny nose”, “sneezing”, “blocked nose”, “unusual eye soreness or discomfort”, “loss of sense of smell”, “loss of sense of taste”, “sore or painful throat”, “hoarse voice”, “headache”, “dizziness, light-headedness or vertigo”, “shortness of breath or trouble breathing affecting normal activities”, “new persistent cough”, “tightness in the chest”, “chest pain”, “racing heart or palpitations”, “fever”, “chills (feeling too cold)”, “difficulty sleeping”, “felt more tired than normal”, “severe fatigue”, “numbness or tingling somewhere in the body”, “feeling of heaviness in arms or legs”, “strong muscle pains or aches”, “shaking or difficulty while walking”, “phlegm production/chesty cough”, “raised, red, itchy welts on the skin or sudden swelling of the face or lips”, “red/purple sores or blisters on feet”, “confusion, disorientation or drowsiness”) and were asked to report whether they had experienced the listed symptom during the months of February-March 2020; April-May 2020;
June-July 2020 (July questionnaire) and / or July-August 2020; September-October 2020 (November questionnaire).

Only 28 of the 33 symptoms were included for consideration as five symptoms (“runny nose”, “sneezing”, “blocked nose”, “shaking or difficulty while walking” and “phlegm production/chesty cough”) were considered non-specific.

Similarly to BiB, data were used to derive the symptom length categories above through summing whether any of the included symptoms (i.e. were present for 0-4 weeks; 4-12 weeks or 12+ weeks, at any point in time over the specified period. This was performed for people who had had COVID-19 and those who had not (confirmed by negative antibody testing).

**Supplementary Information 3:** Detail of method to derive inverse probability weights (IPW)

Self-reported COVID-19 status was regressed on each exposure to assess whether COVID-19 was associated with each socio-demographic or pre-pandemic health risk factor. To determine what variables to include across LS, observed associations were meta-analysed to identify consistent predictors of COVID-19 self-report status (see Supplementary Information 2 for list of covariates used to derive IPWs). To avoid missingness on IPWs, covariates included in each model were imputed using multiple imputation by chained equations (MICE) and IPWs were derived across multiple imputed data sets. All statistical analyses on the LS were performed in Stata version 16 or R (release 3.6.0 or later).

Covariates included in all longitudinal studies

•	Sex

•	Age

•	Ethnicity

•	Mental Health Score

•	BMI

Additional covariates included in some longitudinal studies

•	Asthma (NCDS)

•	Smoking (NCDS; BCS70; MCS; NS)
